# Data-driven profiling of accelerometer-determined physical behaviors and health outcomes in adults: A systematic review

**DOI:** 10.1101/2025.08.29.25334724

**Authors:** Vahid Farrahi, Esmaeil Farhang

## Abstract

**Objective:** Data-driven population segmentation analysis are analytical approaches applicable for profiling physical behaviors from wearable accelerometry data. These methods rely on data rather than predefined, knowledge-driven hypotheses to first identify multiple subgroups within a sample population and then evaluate their relationships with health outcomes. This systematic review aims to describe and synthesize multidimensional physical behavior profiles derived from accelerometry data across adult populations, as identified using data-driven population segmentation analysis.

**Method:** Three electronic databases were searched for relevant articles published up to July 2025. Peer-reviewed journal articles that applied data-driven population segmentation analysis to accelerometer-monitored physical behaviors in adult participants (>18 years) to create profiles of physical behaviors, and that examined the associations of the profiles with a health outcome, were considered. Studies conducted with clinical or specific sample populations were not considered.

**Results:** Of the 16,289 publications retrieved, 40 were included. The most commonly employed technique for physical behavior profiling was the machine learning K-means clustering algorithm (n=18), followed by latent profile analysis (n=8). A diverse set of descriptor variables was derived from accelerometer signals and utilized. The review of profiles of physical behavior revealed several hypothesis-generating, preliminary evidence about how different components and/or aspects of physical behaviors could cluster together and influence health outcomes.

**Conclusion:** Data-driven population segmentation analysis are viable analytical approaches increasingly employed in accelerometry studies to drive physical behavior profiles. The application of these techniques to accelerometer-measured physical behaviors has generated data-driven findings regarding how various physical behavior profiles may differentially relate to health outcomes.

## INTRODUCTION

Over the past few decades, cohort and epidemiological studies have increasingly relied on wearable accelerometers for measurement of physical activity (PA), sedentary behavior, and sleep, collectively known as physical behaviors (Evenson et al., 2022; Migueles et al., 2022). Modern wearable accelerometers can nowadays capture acceleration signals over multiple days, enabling the quantification of various descriptors that reflect the amount, accumulation patterns, timing, and inter-day and intra-day variations of physical behaviors, among others (Backes et al., 2022; Giurgiu et al., 2025). This enhanced ability to measure physical behaviors in greater detail raises questions about how to best model and interpret the relationships between physical behaviors and health outcomes (Farrahi & Rostami, 2024; Migueles et al., 2022; Pedišić et al., 2017; Rosenberger et al., 2019).

Currently, emerging evidence highlights that physical activities, sedentary behaviors, and sleep do not independently influence health outcomes; rather, these behaviors often interact, cluster together, and exhibit codependent relationships with various health markers and indicators (Chastin, Palarea-Albaladejo, et al., 2015; Migueles et al., 2022; Pedišić, 2014; Pedišić et al., 2017). Further complicating this, accumulating evidence suggests that the timing of PA (H. Feng et al., 2023), the regularity and stability of the rest-activity rhythmicity (Makarem et al., 2024), and patterns of accumulation of sedentary and activity behaviors (Herold et al., 2025) are also important characteristics of one’s physical behavior profile that may be linked to health outcomes. Given these complexities, proper modeling of the relationships between accelerometer-measured physical behaviors and health outcomes requires careful consideration of the analytical approaches and their adequacy for exploring such potentially sophisticated relationships (Migueles et al., 2022; Pedišić, 2014; Pedišić et al., 2017).

Historically, the dominant approach for exploring the relationships between measures of physical behaviors and health outcomes was to employ classical regression approaches (Pedišić, 2014; Pedišić et al., 2017). Recently, new evidence has highlighted a complex interplay between different components and dimensions of physical behaviors (Rollo et al., 2020; Rosenberger et al., 2019), leading to the development of innovative analytical techniques. Analytical approaches, such as compositional data analysis (Dumuid et al., 2019; McGregor et al., 2020), isotemporal substitution methods (Mekary et al., 2009; Migueles et al., 2022), multivariate pattern analysis (Aadland et al., 2019), and functional data analysis (Goldsmith et al., 2015) are now frequently used for exploring the health impacts of physical behaviors (Blodgett et al., 2024; Farrahi et al., 2023; Leviäkangas et al., 2024; Niemelä et al., 2023; Seppänen et al., 2025). These novel techniques are generally known to have improved capacity for better modeling of the complex relationships between PA, sedentary behaviors, and sleep with health outcomes over classical statistical approaches (Migueles et al., 2022). From a methodological perspective, however, such analytical approaches can be considered as variable-centered methods. A variable-centered method refers to techniques that focus on exploring the relationships between one or more variables of interest and an outcome variable (Howard & Hoffman, 2017; Woo et al., 2024), assessing how changes in one or more independent variables (e.g., physical activity, sedentary behavior, sleep) impact the outcome variable (e.g., weight).

Another type of analytical approach often used for modeling the health influence of accelerometer-measured physical behaviors is cluster analysis (Kebede et al., 2024; Migueles et al., 2022). Cluster analysis divide the study population into distinct smaller groups typically based on a diverse set of descriptive variables (Ezugwu et al., 2022; Howard & Hoffman, 2017; Liu et al., 2023; Weller et al., 2020; Woo et al., 2024; Xu & Tian, 2015) and are often referred to as data-driven population segmentation analysis when applied to individual-level data and characteristics (i.e., to distinguish it from applications such as image clustering and segmentation) (Liu et al., 2023; Yan et al., 2018). From a methodological perspective, data-driven population segmentation analysis are conceptually different from variable-centered approaches (Woo et al., 2024). Rather than focusing on relationships among variables, these methods use input descriptors to form multiple subgroups in a data-oriented manner, maximizing within-group similarity and between-group dissimilarity based on individual characteristics (Howard & Hoffman, 2017; Liu et al., 2023). Such methods are considered data-driven approach because the subgroups are formed solely based on input descriptor variables, with subsequent analysis focusing on understanding the differential associations between these derived subgroups and health outcomes. In practice, several clustering algorithms are used for data-driven population segmentation analyses (Ezugwu et al., 2022; Woo et al., 2024; Xu & Tian, 2015), with k-means clustering, hierarchical clustering, and latent profile analysis being among the most commonly applied methods in health and medical research (Liu et al., 2023; Yan et al., 2018).

When applied to characteristics of physical behaviors, the subgroups identified and formed using data-driven population segmentation analysis can be referred to as physical behavior profiles (Kebede et al., 2024). These profiles reflect various behavioral patterns within the sample population. Profiles of individuals grouped by similar physical behaviors can then be compared with other profiles in terms of how the variables combine to create these profiles and how these combinations relate differentially to health outcomes. Despite the growing reliance on data-driven population segmentation analysis to derive multidimensional profiles of physical behaviors from wearable accelerometry data across various populations, it remains unclear what types of profiles would emerge in different adult populations and how these might relate to health outcomes. Additionally, the potential of data-driven population segmentation analysis as an analytical approach for exploring the health associations in physical behavior research needs further clarification.

This study sought to systematically review the studies that created and identified profiles of physical behaviors among adult participants with sample populations that were not restricted to specific health conditions, using data-driven population segmentation analysis (i.e., clustering analysis) and wearable accelerometry data. The objectives of this review were to (1) identify and describe physical behavior profiles emerged in studies applying data-driven methods on wearable accelerometer-determined physical behaviors and (2) to examine whether and how these physical behavior profiles relate differentially to health outcomes.

## MATERIALS AND METHODS

This systematic review was registered with the International Prospective Register of Systematic Reviews (ID: CRD42024612460) and performed in accordance with the Preferred Reporting Items for Systematic Reviews and Meta-Analyses (PRISMA) guidelines (Page et al., 2021).

### Search strategy

Three electronic databases (PubMed, Scopus, and Web of Science) were used to identify relevant articles. The initial search was conducted in June, 2024 and repeated in July 2025. The search strings were reported in the Supplementary File. The search results were limited for human subjects and English-language articles. These were not limited by date and included papers from all years. Additional articles were identified by searching the references of relevant articles identified by the search strategy.

### Eligibility criteria

Full-text original peer-reviewed journal articles applying data-driven population segmentation analysis to accelerometer-monitored physical behaviors in adult participants (18 years or older) to create profiles of physical behaviors and examining the associations of the formed profiles with a health outcome were considered. We defined “health outcome” as any health-related problem that could be influenced by components (PA intensities, sedentary behaviors, and sleep) or any of the dimensions (e.g., timing, temporal distribution, accumulation patterns, or daily/weekly frequencies, etc.) of physical behaviors. Given that data-driven approaches can be used for different purposes on the same dataset, studies performed with the same analytical sample were included.

Studies were excluded if they met at least one of the following criteria: (1) physical behaviors were not determined using wearable accelerometers; (2) data-driven profiles were created by combining measures of accelerometer-determined physical behaviors with nonphysical behavior measures (e.g., diet, background information, etc.); (3) profiles were formed based on knowledge-driven criteria without employing a data-driven approach (e.g., (Länsitie et al., 2021)); (4) the associations between identified physical behavior profiles and health outcomes were not statistically examined; (5) the study was not conducted with adult participants; or (6) the study was conducted with clinical or other specific populations.

### Study selection

Covidence review management software was used for the screening and selection of records retrieved from the databases. One person (VF) performed the initial search and screened the titles and abstracts of all articles identified by the search strings to exclude articles that were not relevant according to the eligibility criteria. Articles identified in the title and abstract were first assessed for eligibility and then examined in full text independently by the authors (VF and EF). The reviewers were not blinded to authors or journals. Conflicting decisions were resolved by discussion between the authors until full consensus was achieved. Reasons for exclusion of articles after full-article screening were recorded. Included articles were then reviewed and data from the studies were extracted by one of the authors (VF).

### Data extraction

The extracted data items on the characteristics of the included studies were first author, year of publication, sample population, country, study design (cross-sectional or prospective design), number of participants included in profile analyses, age of participants, proportion of women, and the primary health outcomes. For profile analysis, extracted data items included input descriptor variables used for profile analysis and the method used for driving the profiles. The identified profiles and the number of participants within each profile were recorded. Statistically significant associations observed between physical behavior profiles and health outcomes were also recorded. Data items related to accelerometer data collection methods and characteristics included the type of device used (uniaxial or triaxial), device placement, wear time protocol, minimum valid hours and days required for inclusion in analyses, and criteria for nonwear time.

### Assessment of bias

Risk of bias was assessed using a combination of two tools, adapted to accommodate both the observational nature of the studies and the wearable-accelerometer measurements. Four items from the Newcastle–Ottawa Quality Assessment Scale (Wells et al., 2000) and eight items based on recommended reporting practices for accelerometer studies were utilized (Montoye et al., 2018). These items evaluated the quality of selection strategies, outcome measures, and reporting methods related to accelerometers. A total quality score was determined, with a maximum of 11 points for cross-sectional studies and 12 points for prospective cohort studies (low quality [1–5 points], moderate quality [6–10 points], and high quality [10–11/12 points]).

### Synthesis of results

The derived profiles could be largely attributed to the input descriptor variables used for profile analysis. For better synthesis of the results regarding the health implications of identified profiles, the publications were categorized into five groups based on the input descriptors and types of profiles identified in each study. These categories were PA profiles, sedentary behavior profiles, sleep profiles, rest-activity profiles, and joint profiles. Joint profiles refer to those profiles that were created by incorporating descriptors from at least two components of physical behaviors (i.e., any combination of descriptors of PA, sedentary behavior, and sleep).

We presented the overview of profiles in color-coded figures. The input descriptors used for profile analysis, the derived profiles, statistically significant differences in health outcomes among profiles, and statistical modeling used for examining the associations were presented in tables. A persistent challenge in wearable accelerometer-based measurement is the incomparability of study results because of large variability in decisions related to accelerometer data cleaning and processing (Pedišić & Bauman, 2015). To enable a better comparison among physical behavior profiles identified, profiles with comparable overall characteristics that were seen in *two or more distinct* sample populations were identified and marked.

The common approach to examining health associations with data-driven profiles involves assessing differences relative to a referent profile. When selecting a reference profile, it is typically chosen based on its representation of either the unhealthiest or healthiest physical behavior profile. Comparing the differences in the health outcomes among the derived profiles with respect to one another could also be performed. When statistically significant associations or differences were observed between a profile and a health outcome, both the direction of the association and the referent group were indicated. Differences were denoted using upward arrows (↑) to indicate higher values in the health outcome relative to the referent group. Downward arrows (↓) were used to indicate lower values. Only statistically significant associations were reported. For analyses that were gender-stratified, statistical significance was only noted if the observed associations were significant for both men and women. If multiple unadjusted and covariate-adjusted models were built, the significant association from the fully adjusted models was extracted and reported.

The characteristics and health associations of profiles identified across two or more sample populations were compared and synthesized narratively. Other key emerging profiles in the studies, together with the health associations linked to the key characteristics, were also described and summarized narratively.

## RESULTS

A flow diagram of the included publications at each stage is presented in Figure 1. Of the 16,289 records initially retrieved (3,244 from PubMed, 7,390 from Scopus, and 5,655 from Web of Science), 7,156 duplicates were removed. Following the screening of titles and abstracts, 60 articles were identified as potentially relevant. After conducting a full-text review of these 60 publications, 38 were deemed eligible for inclusion. Additionally, two more articles were identified through hand/citation searches, resulting in a final total of 40 included studies.

**Figure 1:**
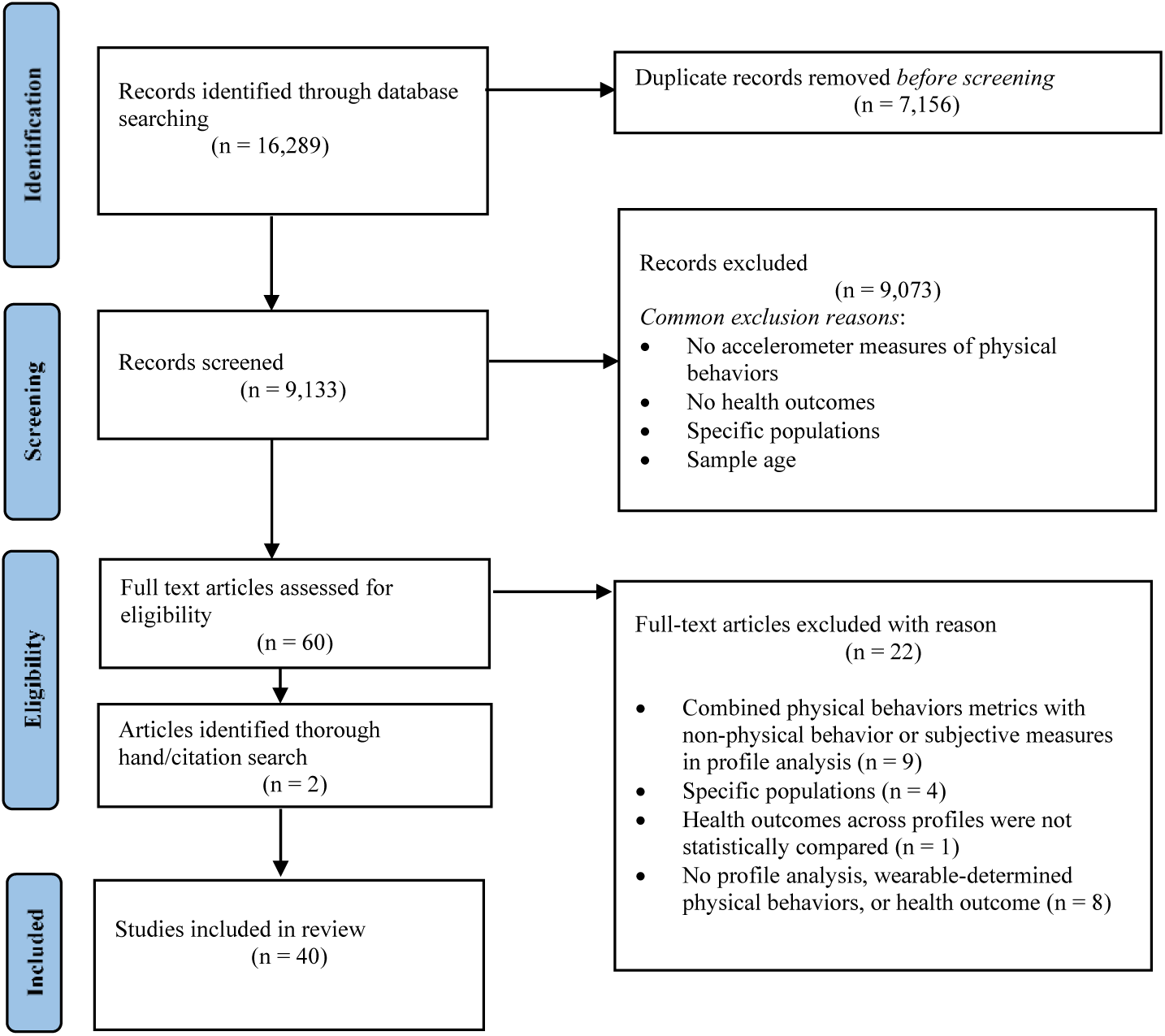
Flow diagram showing the selection of studies for inclusion in the systematic review

### Study design, participant characteristics, and publication characteristics

The main characteristics of the included studies are summarized in Table 1. Publication dates for the 40 included articles ranged from 2013 to 2025, with the majority published since 2019 (n = 36). Of these studies, 25 were conducted using a cross-sectional design (Aqeel et al., 2021; Biswas et al., 2022, 2023; Choi et al., 2025; Farrahi et al., 2021, 2022, 2024; Fukuoka et al., 2018; Full et al., 2019; Graves et al., 2021; Gupta et al., 2020, 2025; Hibbing et al., 2022; P. H. Lee et al., 2013; Leskinen et al., 2023; Ma et al., 2023; Marschollek, 2016; Matricciani et al., 2021; Nawrin et al., 2024; Niemelä et al., 2019; O’Regan et al., 2021; Palmberg et al., 2024; Paolillo et al., 2023; Smagula et al., 2022; Yamamoto et al., 2023) while 15 studies had a prospective study design (Albalak et al., 2023; Bai, Ning, et al., 2024; Bai, Shao, et al., 2024; Bai, Zhou, et al., 2024; Chung et al., 2024; Evenson et al., 2017; Fabrizio et al., 2024; Q. Feng et al., 2025; Li et al., 2024; Reuter et al., 2020; Shim et al., 2023; von Rosen et al., 2020; Yamamoto et al., 2025; Yerramalla et al., 2024; Zhang et al., 2024).

**Table 1:**
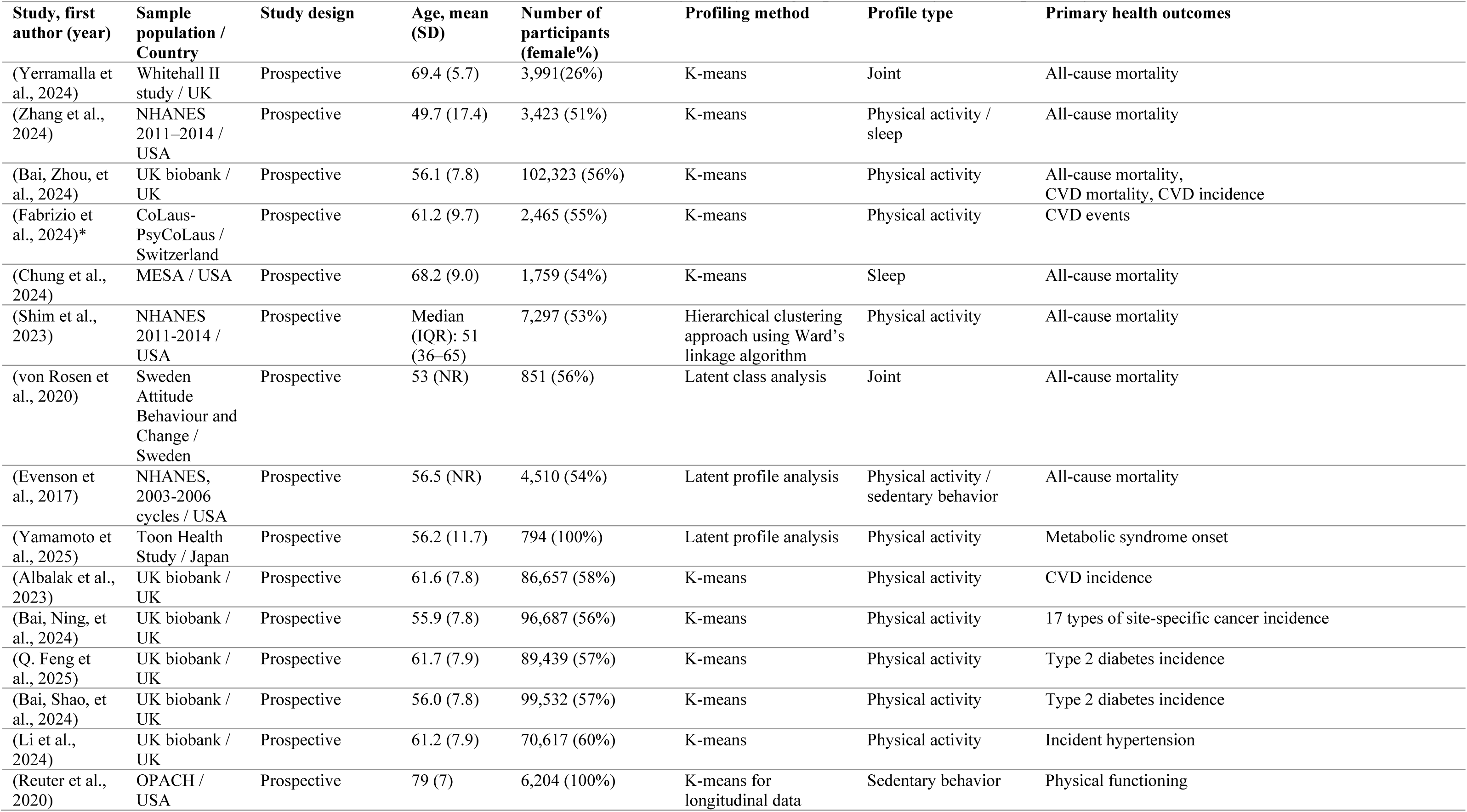
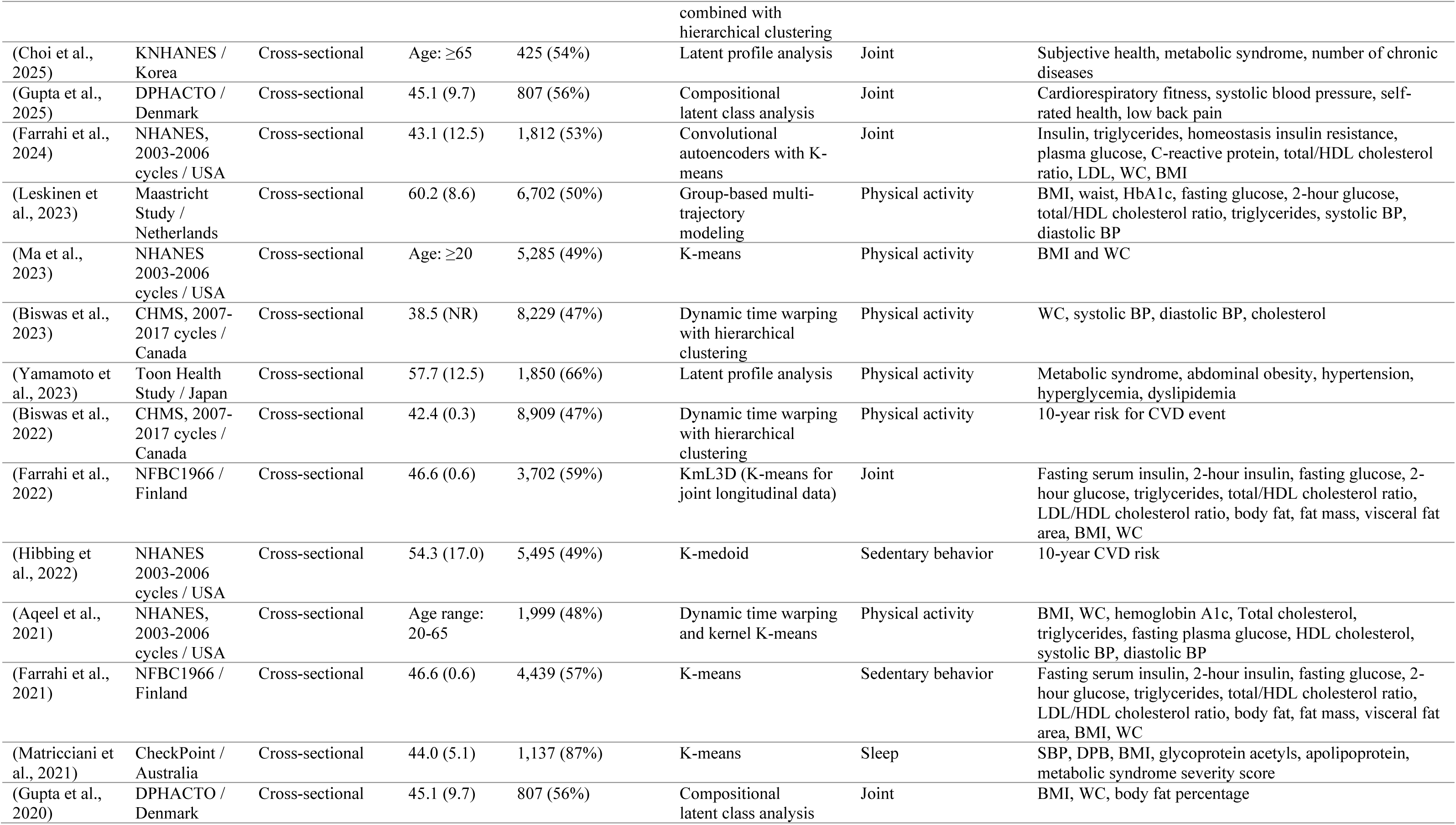
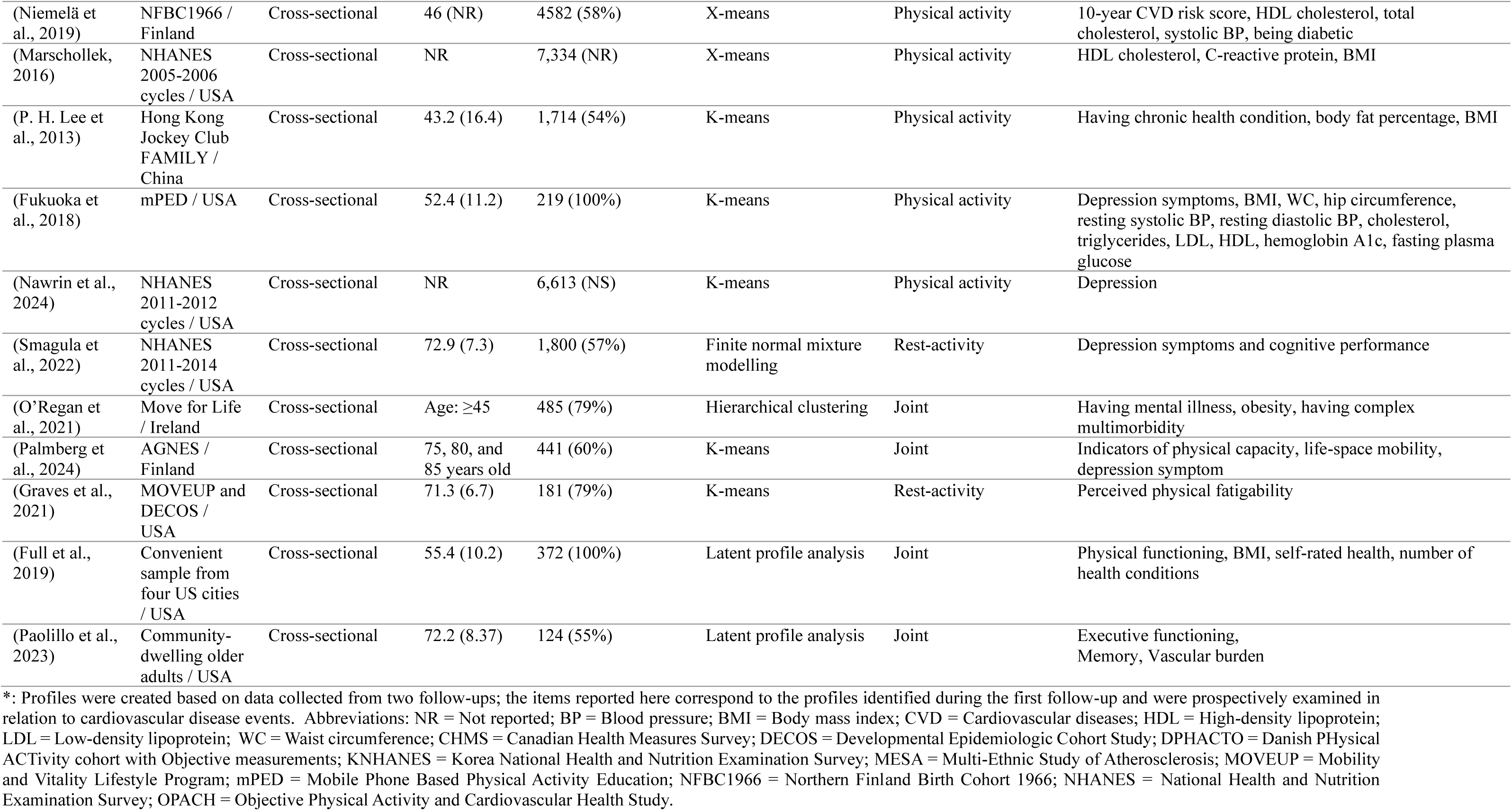
Main characteristics of 40 studies included in the review, listed by study design, publication year, and primary health outcome.

| Study, first author (year) | Sample population / Country | Study design | Age, mean (SD) | Number of participants (female%) | Profiling method | Profile type | Primary health outcomes |
| --- | --- | --- | --- | --- | --- | --- | --- |
| (Yerramalla et al., 2024) | Whitehall II study / UK | Prospective | 69.4 (5.7) | 3,991 (26%) | K-means | Joint | All-cause mortality |
| (Zhang et al., 2024) | NHANES 2011–2014 / USA | Prospective | 49.7 (17.4) | 3,423 (51%) | K-means | Physical activity / sleep | All-cause mortality |
| (Bai, Zhou, et al., 2024) | UK biobank / UK | Prospective | 56.1 (7.8) | 102,323 (56%) | K-means | Physical activity | All-cause mortality, CVD mortality, CVD incidence |
| (Fabrizio et al., 2024)* | CoLaus-PsyCoLaus / Switzerland | Prospective | 61.2 (9.7) | 2,465 (55%) | K-means | Physical activity | CVD events |
| (Chung et al., 2024) | MESA / USA | Prospective | 68.2 (9.0) | 1,759 (54%) | K-means | Sleep | All-cause mortality |
| (Shim et al., 2023) | NHANES 2011–2014 / USA | Prospective | Median (IQR): 51 (36–65) | 7,297 (53%) | Hierarchical clustering approach using Ward’s linkage algorithm | Physical activity | All-cause mortality |
| (von Rosen et al., 2020) | Sweden Attitude Behaviour and Change / Sweden | Prospective | 53 (NR) | 851 (56%) | Latent class analysis | Joint | All-cause mortality |
| (Evenson et al., 2017) | NHANES, 2003–2006 cycles / USA | Prospective | 56.5 (NR) | 4,510 (54%) | Latent profile analysis | Physical activity / sedentary behavior | All-cause mortality |
| (Yamamoto et al., 2025) | Toon Health Study / Japan | Prospective | 56.2 (11.7) | 794 (100%) | Latent profile analysis | Physical activity | Metabolic syndrome onset |
| (Albalak et al., 2023) | UK biobank / UK | Prospective | 61.6 (7.8) | 86,657 (58%) | K-means | Physical activity | CVD incidence |
| (Bai, Ning, et al., 2024) | UK biobank / UK | Prospective | 55.9 (7.8) | 96,687 (56%) | K-means | Physical activity | 17 types of site-specific cancer incidence |
| (Q. Feng et al., 2025) | UK biobank / UK | Prospective | 61.7 (7.9) | 89,439 (57%) | K-means | Physical activity | Type 2 diabetes incidence |
| (Bai, Shao, et al., 2024) | UK biobank / UK | Prospective | 56.0 (7.8) | 99,532 (57%) | K-means | Physical activity | Type 2 diabetes incidence |
| (Li et al., 2024) | UK biobank / UK | Prospective | 61.2 (7.9) | 70,617 (60%) | K-means | Physical activity | Incident hypertension |
| (Reuter et al., 2020) | OPACH / USA | Prospective | 79 (7) | 6,204 (100%) | K-means for longitudinal data combined with<br>hierarchical clustering | Sedentary behavior | Physical functioning |
| (Choi et al., 2025) | KNHANES / Korea | Cross-sectional | Age: $\geq 65$ | 425 (54%) | Latent profile analysis | Joint | Subjective health, metabolic syndrome, number of chronic diseases |
| (Gupta et al., 2025) | DPHACTO / Denmark | Cross-sectional | 45.1 (9.7) | 807 (56%) | Compositional latent class analysis | Joint | Cardiorespiratory fitness, systolic blood pressure, self-rated health, low back pain |
| (Farrahi et al., 2024) | NHANES, 2003-2006 cycles / USA | Cross-sectional | 43.1 (12.5) | 1,812 (53%) | Convolutional autoencoders with K-means | Joint | Insulin, triglycerides, homeostasis insulin resistance, plasma glucose, C-reactive protein, total/HDL cholesterol ratio, LDL, WC, BMI |
| (Leskinen et al., 2023) | Maastricht Study / Netherlands | Cross-sectional | 60.2 (8.6) | 6,702 (50%) | Group-based multi-trajectory modeling | Physical activity | BMI, waist, HbA1c, fasting glucose, 2-hour glucose, total/HDL cholesterol ratio, triglycerides, systolic BP, diastolic BP |
| (Ma et al., 2023) | NHANES 2003-2006 cycles / USA | Cross-sectional | Age: $\geq 20$ | 5,285 (49%) | K-means | Physical activity | BMI and WC |
| (Biswas et al., 2023) | CHMS, 2007-2017 cycles / Canada | Cross-sectional | 38.5 (NR) | 8,229 (47%) | Dynamic time warping with hierarchical clustering | Physical activity | WC, systolic BP, diastolic BP, cholesterol |
| (Yamamoto et al., 2023) | Toon Health Study / Japan | Cross-sectional | 57.7 (12.5) | 1,850 (66%) | Latent profile analysis | Physical activity | Metabolic syndrome, abdominal obesity, hypertension, hyperglycemia, dyslipidemia |
| (Biswas et al., 2022) | CHMS, 2007-2017 cycles / Canada | Cross-sectional | 42.4 (0.3) | 8,909 (47%) | Dynamic time warping with hierarchical clustering | Physical activity | 10-year risk for CVD event |
| (Farrahi et al., 2022) | NFBC1966 / Finland | Cross-sectional | 46.6 (0.6) | 3,702 (59%) | KmL3D (K-means for joint longitudinal data) | Joint | Fasting serum insulin, 2-hour insulin, fasting glucose, 2-hour glucose, triglycerides, total/HDL cholesterol ratio, LDL/HDL cholesterol ratio, body fat, fat mass, visceral fat area, BMI, WC |
| (Hibbing et al., 2022) | NHANES 2003-2006 cycles / USA | Cross-sectional | 54.3 (17.0) | 5,495 (49%) | K-medoid | Sedentary behavior | 10-year CVD risk |
| (Aqeel et al., 2021) | NHANES, 2003-2006 cycles / USA | Cross-sectional | Age range: 20-65 | 1,999 (48%) | Dynamic time warping and kernel K-means | Physical activity | BMI, WC, hemoglobin A1c, Total cholesterol, triglycerides, fasting plasma glucose, HDL cholesterol, systolic BP, diastolic BP |
| (Farrahi et al., 2021) | NFBC1966 / Finland | Cross-sectional | 46.6 (0.6) | 4,439 (57%) | K-means | Sedentary behavior | Fasting serum insulin, 2-hour insulin, fasting glucose, 2-hour glucose, triglycerides, total/HDL cholesterol ratio, LDL/HDL cholesterol ratio, body fat, fat mass, visceral fat area, BMI, WC |
| (Matricciani et al., 2021) | CheckPoint / Australia | Cross-sectional | 44.0 (5.1) | 1,137 (87%) | K-means | Sleep | SBP, DPB, BMI, glycoprotein acetyls, apolipoprotein, metabolic syndrome severity score |
| (Gupta et al., 2020) | DPHACTO / Denmark | Cross-sectional | 45.1 (9.7) | 807 (56%) | Compositional latent class analysis | Joint | BMI, WC, body fat percentage |
| (Niemelä et al., 2019) | NFBC1966 / Finland | Cross-sectional | 46 (NR) | 4582 (58%) | X-means | Physical activity | 10-year CVD risk score, HDL cholesterol, total cholesterol, systolic BP, being diabetic |
| (Marschollek, 2016) | NHANES 2005-2006 cycles / USA | Cross-sectional | NR | 7,334 (NR) | X-means | Physical activity | HDL cholesterol, C-reactive protein, BMI |
| (P. H. Lee et al., 2013) | Hong Kong Jockey Club FAMILY / China | Cross-sectional | 43.2 (16.4) | 1,714 (54%) | K-means | Physical activity | Having chronic health condition, body fat percentage, BMI |
| (Fukuoka et al., 2018) | mPED / USA | Cross-sectional | 52.4 (11.2) | 219 (100%) | K-means | Physical activity | Depression symptoms, BMI, WC, hip circumference, resting systolic BP, resting diastolic BP, cholesterol, triglycerides, LDL, HDL, hemoglobin A1c, fasting plasma glucose |
| (Nawrin et al., 2024) | NHANES 2011-2012 cycles / USA | Cross-sectional | NR | 6,613 (NS) | K-means | Physical activity | Depression |
| (Smagula et al., 2022) | NHANES 2011-2014 cycles / USA | Cross-sectional | 72.9 (7.3) | 1,800 (57%) | Finite normal mixture modelling | Rest-activity | Depression symptoms and cognitive performance |
| (O'Regan et al., 2021) | Move for Life / Ireland | Cross-sectional | Age: $\geq 45$ | 485 (79%) | Hierarchical clustering | Joint | Having mental illness, obesity, having complex multimorbidity |
| (Palmberg et al., 2024) | AGNES / Finland | Cross-sectional | 75, 80, and 85 years old | 441 (60%) | K-means | Joint | Indicators of physical capacity, life-space mobility, depression symptom |
| (Graves et al., 2021) | MOVEUP and DECOS / USA | Cross-sectional | 71.3 (6.7) | 181 (79%) | K-means | Rest-activity | Perceived physical fatigability |
| (Full et al., 2019) | Convenient sample from four US cities / USA | Cross-sectional | 55.4 (10.2) | 372 (100%) | Latent profile analysis | Joint | Physical functioning, BMI, self-rated health, number of health conditions |
| (Paolillo et al., 2023) | Community-dwelling older adults / USA | Cross-sectional | 72.2 (8.37) | 124 (55%) | Latent profile analysis | Joint | Executive functioning, Memory, Vascular burden |

\*: Profiles were created based on data collected from two follow-ups; the items reported here correspond to the profiles identified during the first follow-up and were prospectively examined in relation to cardiovascular disease events. Abbreviations: NR = Not reported; BP = Blood pressure; BMI = Body mass index; CVD = Cardiovascular diseases; HDL = High-density lipoprotein; LDL = Low-density lipoprotein; WC = Waist circumference; CHMS = Canadian Health Measures Survey; DECOS = Developmental Epidemiologic Cohort Study; DPHACTO = Danish PHysical ACTivity cohort with Objective measurements; KHNHANES = Korea National Health and Nutrition Examination Survey; MESA = Multi-Ethnic Study of Atherosclerosis; MOVEUP = Mobility and Vitality Lifestyle Program; mPED = Mobile Phone Based Physical Activity Education; NFBC1966 = Northern Finland Birth Cohort 1966; NHANES = National Health and Nutrition Examination Survey; OPACH = Objective Physical Activity and Cardiovascular Health Study.

Among all included studies, the number of participants in the sample population used for physical behavior profiling ranged between 124 (Paolillo et al., 2023) to 102,323 (Bai, Zhou, et al., 2024). Sample populations were drawn from 13 countries. Sixteen studies were performed with a sample population from the United States, seven from the United Kingdom, four from Finland, two from Canada, two from Japan, and two from Denmark. Other sample populations were from Sweden, the Netherlands, Switzerland, Ireland, Australia Korea, and China. A large number of studies analyzed accelerometer data from the National Health and Nutrition Examination Survey study (NHANES, n = 10).

### Quality score, data collection methods, and accelerometer characteristics

Details of the accelerometer data collection measures are summarized in Supplementary Table S1. Across the studies, 12 required participants to wear the accelerometer during waking hours for 7 days (Aqeel et al., 2021; Biswas et al., 2022, 2023; Choi et al., 2025; Evenson et al., 2017; Farrahi et al., 2024; Fukuoka et al., 2018; Hibbing et al., 2022; Leskinen et al., 2023; Marschollek, 2016; von Rosen et al., 2020; Yamamoto et al., 2023, 2025), while 11 had continuous 7-day measurement including sleep (Albalak et al., 2023; Bai, Ning, et al., 2024; Bai, Shao, et al., 2024; Bai, Zhou, et al., 2024; Q. Feng et al., 2025; Graves et al., 2021; Li et al., 2024; Nawrin et al., 2024; Reuter et al., 2020; Shim et al., 2023; Zhang et al., 2024). The number of valid accelerometer measurement days deemed sufficient across the studies ranged from 1 to 7 consecutive days.

Individual publication quality scores are shown in Supplementary Table S2. Twelve studies with a prospective study design (Albalak et al., 2023; Bai, Ning, et al., 2024; Bai, Shao, et al., 2024; Evenson et al., 2017; Fabrizio et al., 2024; Q. Feng et al., 2025; Li et al., 2024; Reuter et al., 2020; Shim et al., 2023; von Rosen et al., 2020; Yerramalla et al., 2024; Zhang et al., 2024) and 20 studies with a cross-sectional study design were rated as high quality (Aqeel et al., 2021; Biswas et al., 2022, 2023; Choi et al., 2025; Farrahi et al., 2021, 2022, 2024; Fukuoka et al., 2018; Graves et al., 2021; Gupta et al., 2025, 2020; Hibbing et al., 2022; P. H. Lee et al., 2013; Leskinen et al., 2023; Ma et al., 2023; Nawrin et al., 2024; Niemelä et al., 2019; O’Regan et al., 2021; Palmberg et al., 2024; Yamamoto et al., 2023). Three studies with a prospective study design (Bai, Zhou, et al., 2024; Chung et al., 2024; Yamamoto et al., 2025) and five studies with a cross-sectional study design were rated as moderate quality (Full et al., 2019; Marschollek, 2016; Matricciani et al., 2021; Paolillo et al., 2023; Smagula et al., 2022). No study was rated as low quality.

### Health outcomes

The health outcomes assessed in the included studies are shown in Table 1. In most prospective studies, the primary health outcome (n = 7) was all-cause mortality (Bai, Zhou, et al., 2024; Chung et al., 2024; Evenson et al., 2017; Shim et al., 2023; von Rosen et al., 2020; Yerramalla et al., 2024; Zhang et al., 2024). Other primary health outcomes in prospective studies were cardiovascular disease (CVD) incidence (Albalak et al., 2023; Bai, Zhou, et al., 2024; Fabrizio et al., 2024), type-2 diabetes incidence (Bai, Shao, et al., 2024; Q. Feng et al., 2025), hypertension incidence (Li et al., 2024), onset of metabolic syndrome (Yamamoto et al., 2025), site-specific cancer incidence (Bai, Ning, et al., 2024), and deterioration in physical functioning (Reuter et al., 2020). Primary health outcomes in most studies with a cross-sectional design (n = 16) were indicators of cardiometabolic health, CVD risk factors, metabolic syndrome, and/or obesity indicators (Aqeel et al., 2021; Biswas et al., 2022, 2023; Choi et al., 2025; Farrahi et al., 2021, 2022, 2024; Fukuoka et al., 2018; Gupta et al., 2020; Hibbing et al., 2022; P. H. Lee et al., 2013; Leskinen et al., 2023; Ma et al., 2023; Marschollek, 2016; Matricciani et al., 2021; Niemelä et al., 2019; Yamamoto et al., 2023). Other primary health outcomes in cross-sectional studies were depression or mental health outcomes (Nawrin et al., 2024; O’Regan et al., 2021; Smagula et al., 2022); indicators of physical function capacity, or frailty (Full et al., 2019; Graves et al., 2021; Palmberg et al., 2024); low back pain (Gupta et al., 2025); or indicators of cognitive health (Paolillo et al., 2023).

### Profiling method

Of the 40 included studies (see Table 1), 18 used the K-means clustering algorithm to derive physical behavior profiles (Albalak et al., 2023; Bai, Ning, et al., 2024; Bai, Shao, et al., 2024; Bai, Zhou, et al., 2024; Chung et al., 2024; Fabrizio et al., 2024; Farrahi et al., 2021; Q. Feng et al., 2025; Fukuoka et al., 2018; Graves et al., 2021; P. H. Lee et al., 2013; Li et al., 2024; Ma et al., 2023; Matricciani et al., 2021; Nawrin et al., 2024; Palmberg et al., 2024; Yerramalla et al., 2024; Zhang et al., 2024). Seven of these applied different variations and forms of K-means, including K-means for longitudinal data (Farrahi et al., 2022; Reuter et al., 2020), dynamic time warping and kernel K-means (Aqeel et al., 2021), convolutional autoencoders with K-means (Farrahi et al., 2024), X-means (Marschollek, 2016; Niemelä et al., 2019), and K-medoid (Hibbing et al., 2022). Nine studies used latent profile analysis (Choi et al., 2025; Evenson et al., 2017; Full et al., 2019; Gupta et al., 2020, 2025; Paolillo et al., 2023; von Rosen et al., 2020; Yamamoto et al., 2023, 2025), four employed hierarchical clustering (Biswas et al., 2022, 2023; O’Regan et al., 2021; Shim et al., 2023), one applied finite mixture modelling (Smagula et al., 2022), and one used group-based multi-trajectory modeling (Leskinen et al., 2023).

### Profile types

Among the 40 included studies, 22 identified profiles of PA, four identified profiles of sedentary behavior, three identified profiles of sleep, and two identified profiles of rest-activity rhythmicity. One study reported both PA and sleep profiles (Zhang et al., 2024) and another reported both PA and sedentary profiles (Evenson et al., 2017); these were counted in both respective categories. Joint profiling was performed in 11 studies.

### Profiles of physical activity

An overview of the profiles identified in studies creating PA profiles is shown in Figure 2. The number of profiles identified ranged from two to seven. The input descriptor variables, the derived PA profiles, and their associations with health outcomes in each study are listed in Table 2.

**Figure 2:**
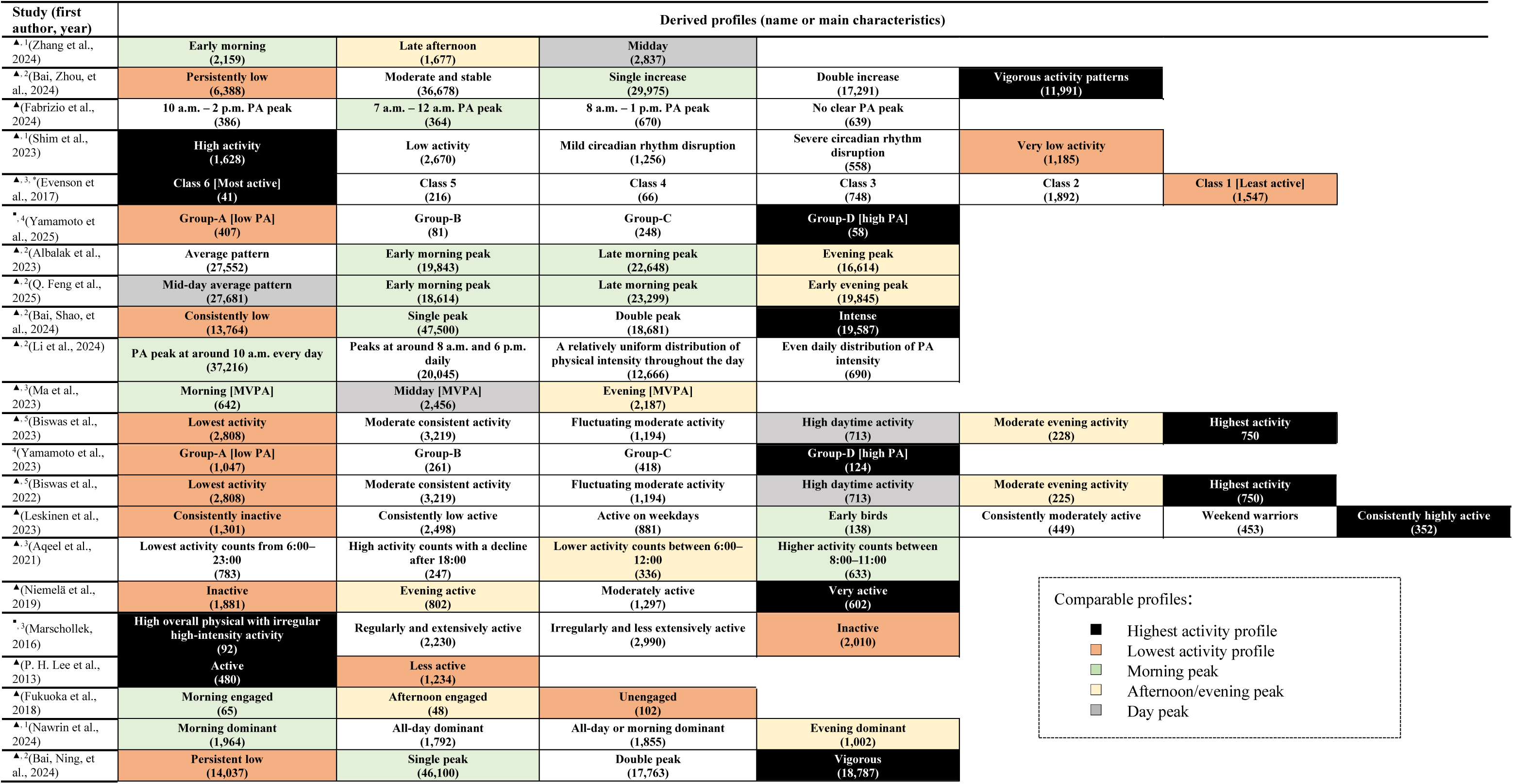
Overview of derived profiles in 22 studied identifying physical activity (PA) profiles. The numbers shown in () are the number of participants in the profile. The profiles are specified according to the names given to them in the study or a short outline of the main characteristics. To facilitate comparison across studies, profiles with comparable overall characteristics *seen in at least two or more distinct sample populations* are highlighted using distinct background colors. Note that the color-coded profiles are not necessarily exactly identical or directly comparable, because of the underlying heterogeneities related to profile analysis and/or accelerometer data cleaning/processing. 1, 2, 3, 4, 5: studies marked with the same number were performed with the same sample population. ▴: High quality study. ▪: Moderate quality study. Individual publication quality scores are shown in Supplementary Table S2. *: Five models were created based on different input descriptor variables into the profiling approach; profiles reported here are for the profiles identified on the basis of average activity counts/minute.

**Table 2:**
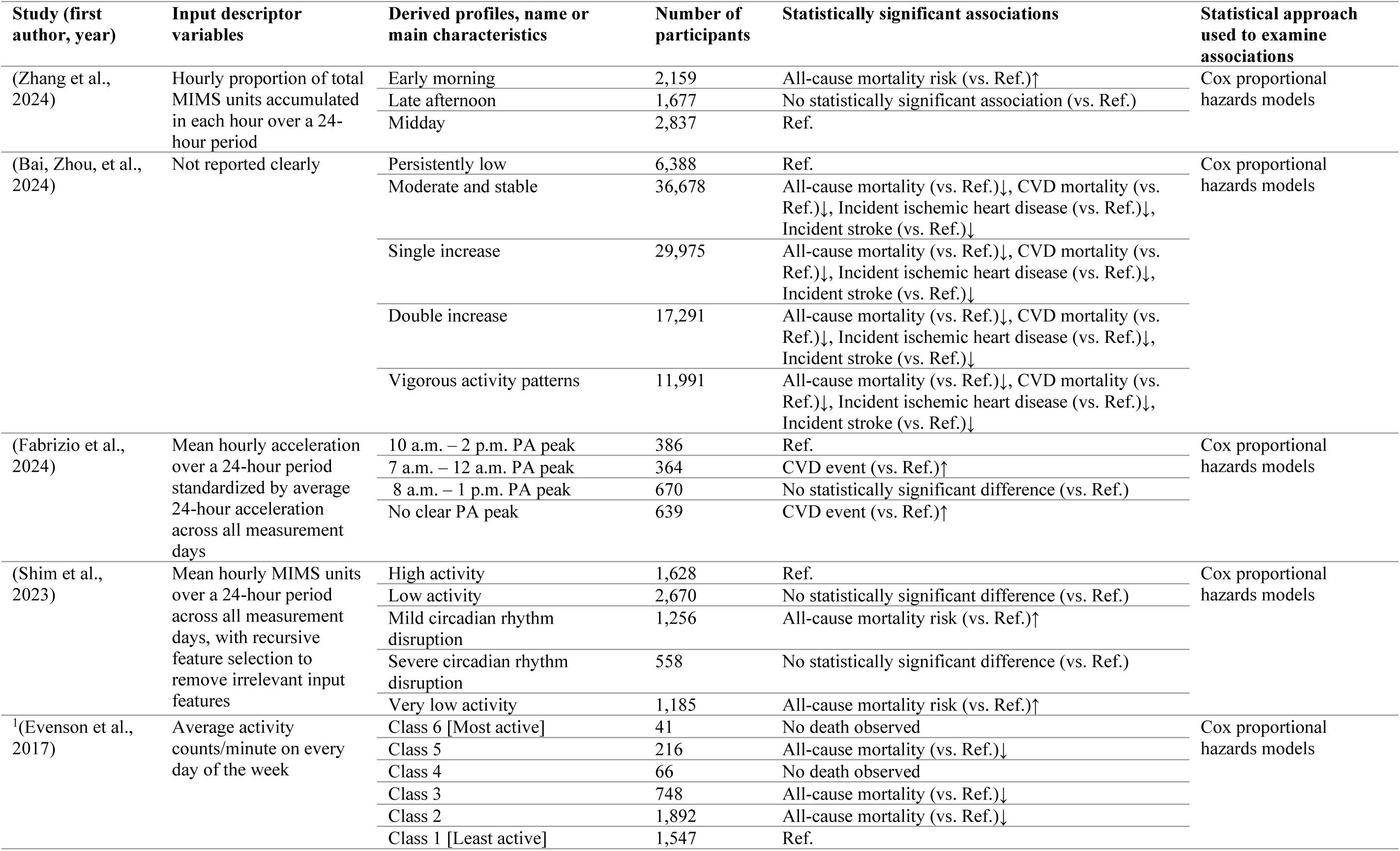
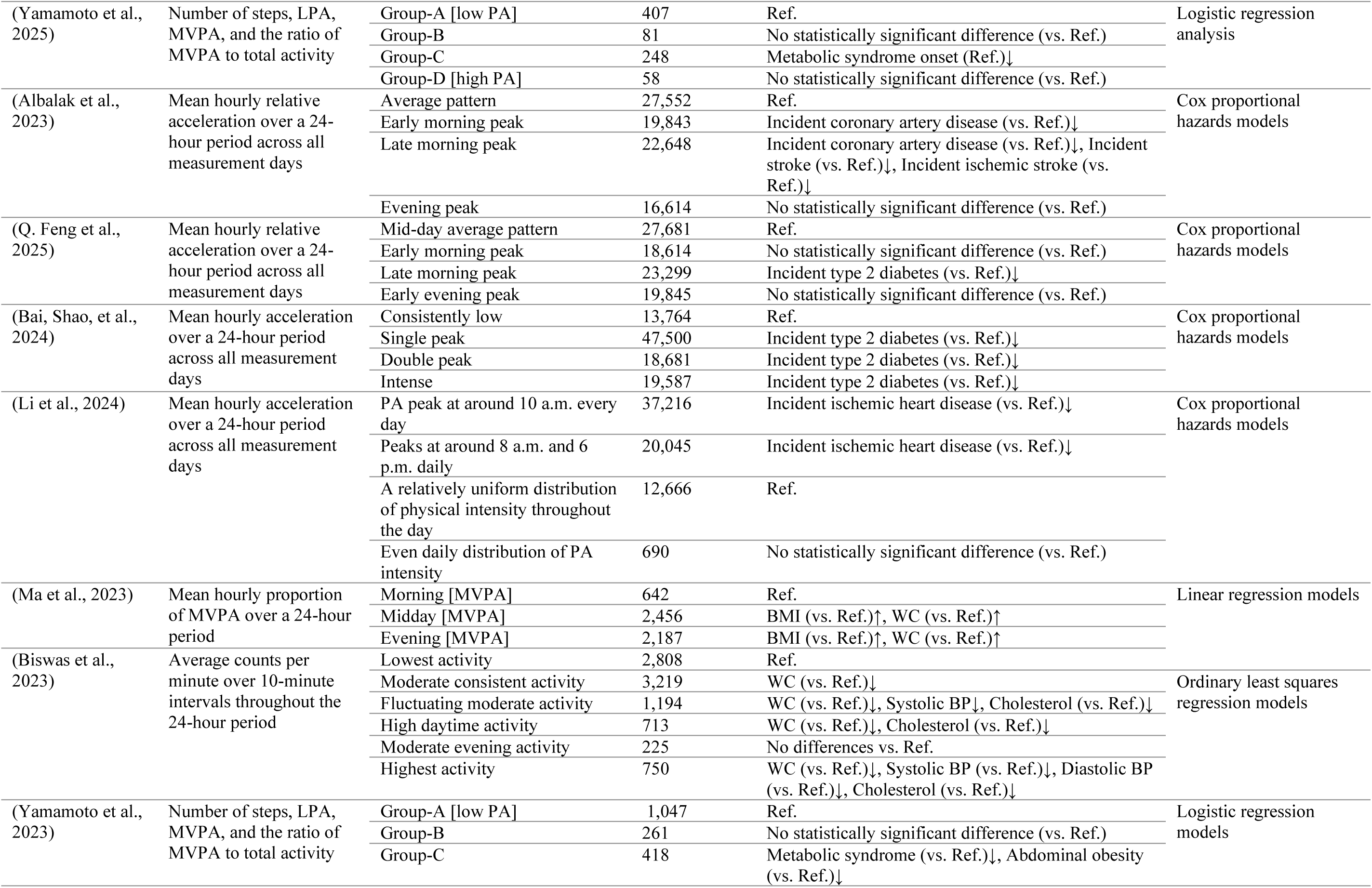
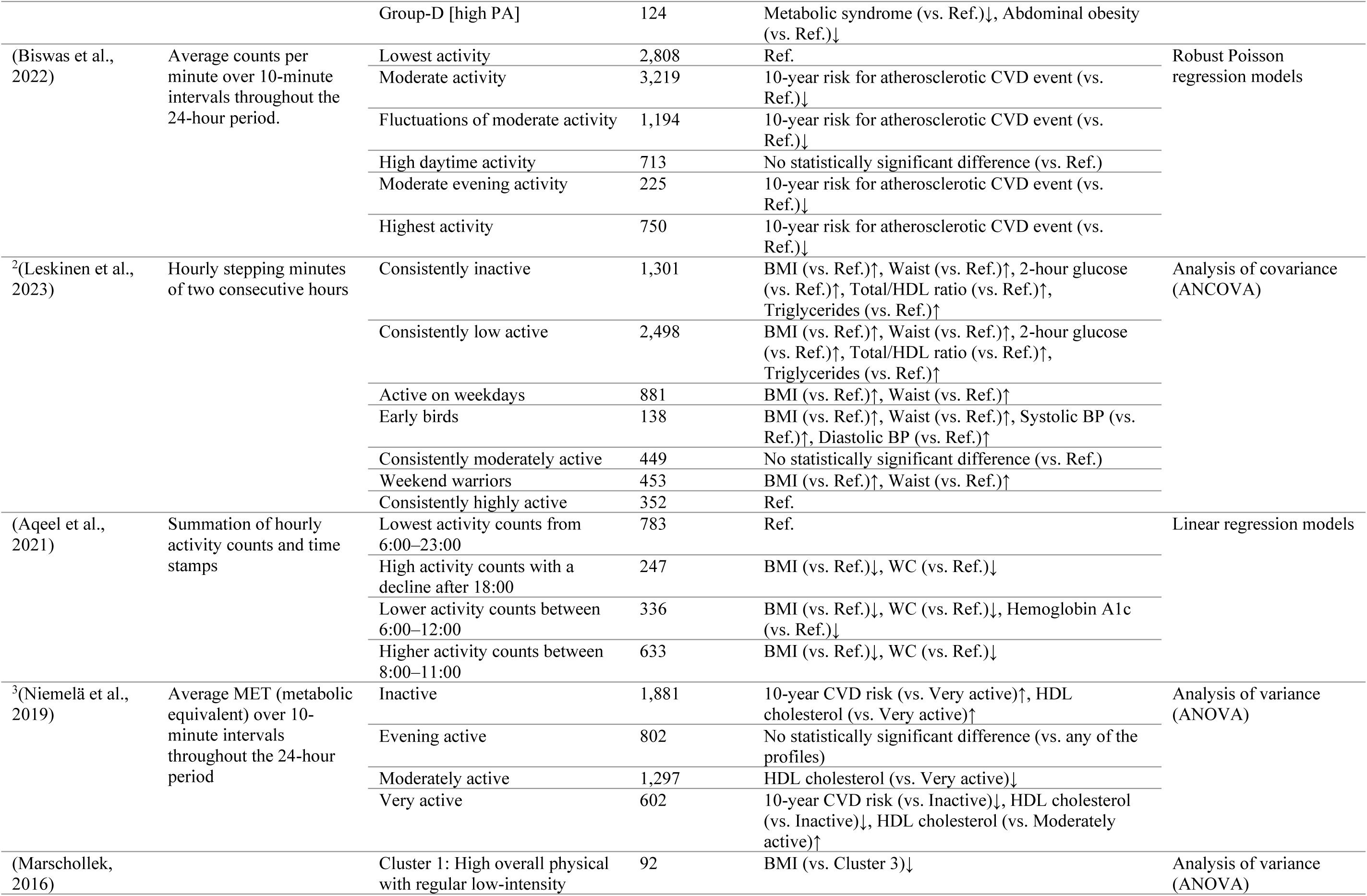
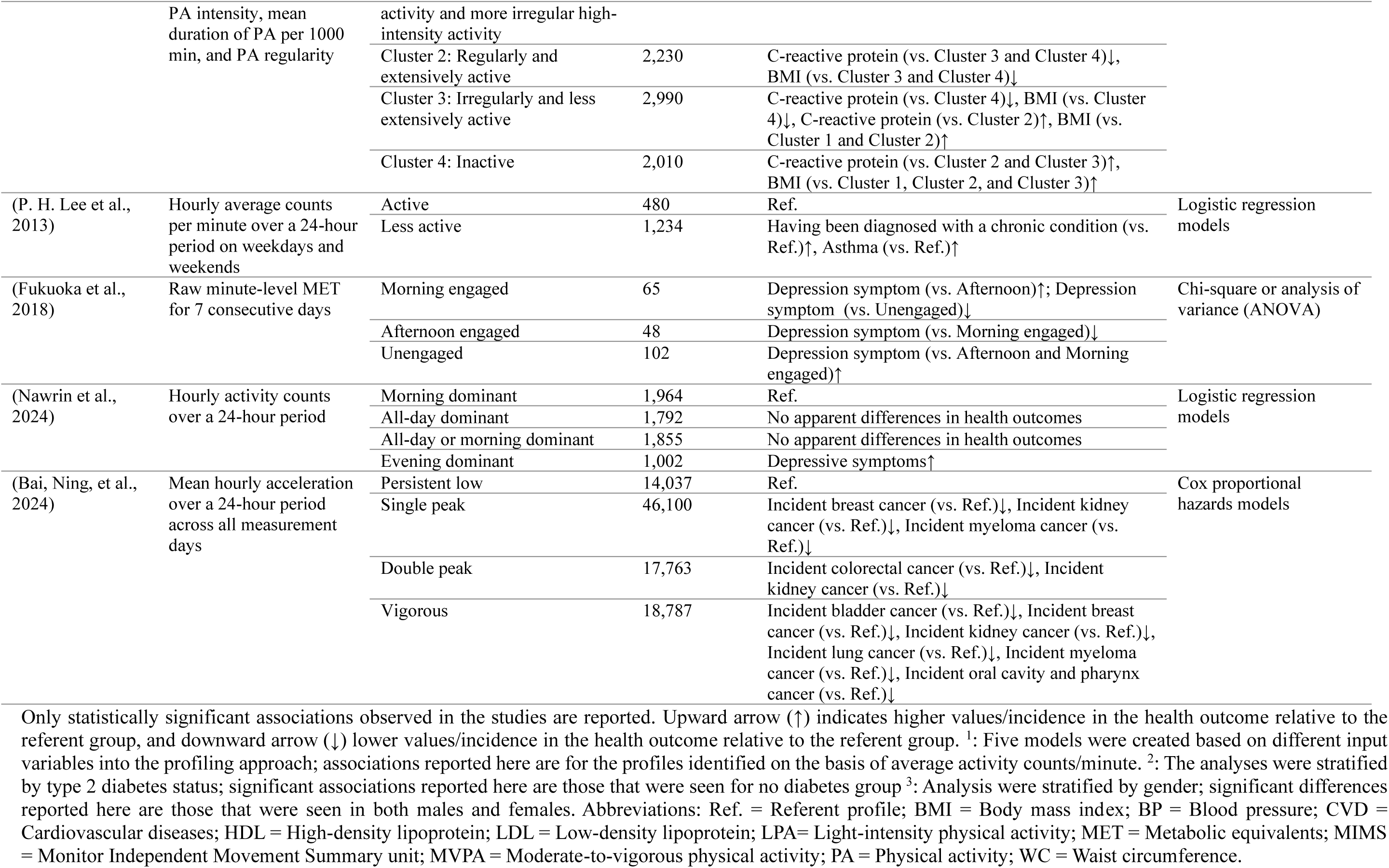
Input descriptor variables, profiles identified, and the statistically significant associations observed among the profiles identified in 22 studies performing physical activity (PA) profiling.

| Study (first author, year) | Input descriptor variables | Derived profiles, name or main characteristics | Number of participants | Statistically significant associations | Statistical approach used to examine associations |
| --- | --- | --- | --- | --- | --- |
| (Zhang et al., 2024) | Hourly proportion of total MIMS units accumulated in each hour over a 24-hour period | Early morning | 2,159 | All-cause mortality risk (vs. Ref.)↑ | Cox proportional hazards models |
|  |  | Late afternoon | 1,677 | No statistically significant association (vs. Ref.) |  |
|  |  | Midday | 2,837 | Ref. |  |
| (Bai, Zhou, et al., 2024) | Not reported clearly | Persistently low | 6,388 | Ref. | Cox proportional hazards models |
|  |  | Moderate and stable | 36,678 | All-cause mortality (vs. Ref.)↓, CVD mortality (vs. Ref.)↓, Incident ischemic heart disease (vs. Ref.)↓, Incident stroke (vs. Ref.)↓ |  |
|  |  | Single increase | 29,975 | All-cause mortality (vs. Ref.)↓, CVD mortality (vs. Ref.)↓, Incident ischemic heart disease (vs. Ref.)↓, Incident stroke (vs. Ref.)↓ |  |
|  |  | Double increase | 17,291 | All-cause mortality (vs. Ref.)↓, CVD mortality (vs. Ref.)↓, Incident ischemic heart disease (vs. Ref.)↓, Incident stroke (vs. Ref.)↓ |  |
|  |  | Vigorous activity patterns | 11,991 | All-cause mortality (vs. Ref.)↓, CVD mortality (vs. Ref.)↓, Incident ischemic heart disease (vs. Ref.)↓, Incident stroke (vs. Ref.)↓ |  |
| (Fabrizio et al., 2024) | Mean hourly acceleration over a 24-hour period standardized by average 24-hour acceleration across all measurement days | 10 a.m. – 2 p.m. PA peak | 386 | Ref. | Cox proportional hazards models |
|  |  | 7 a.m. – 12 a.m. PA peak | 364 | CVD event (vs. Ref.)↑ |  |
|  |  | 8 a.m. – 1 p.m. PA peak | 670 | No statistically significant difference (vs. Ref.) |  |
|  |  | No clear PA peak | 639 | CVD event (vs. Ref.)↑ |  |
| (Shim et al., 2023) | Mean hourly MIMS units over a 24-hour period across all measurement days, with recursive feature selection to remove irrelevant input features | High activity | 1,628 | Ref. | Cox proportional hazards models |
|  |  | Low activity | 2,670 | No statistically significant difference (vs. Ref.) |  |
|  |  | Mild circadian rhythm disruption | 1,256 | All-cause mortality risk (vs. Ref.)↑ |  |
|  |  | Severe circadian rhythm disruption | 558 | No statistically significant difference (vs. Ref.) |  |
|  |  | Very low activity | 1,185 | All-cause mortality risk (vs. Ref.)↑ |  |
| <sup>1</sup>(Evenson et al., 2017) | Average activity counts/minute on every day of the week | Class 6 [Most active] | 41 | No death observed | Cox proportional hazards models |
|  |  | Class 5 | 216 | All-cause mortality (vs. Ref.)↓ |  |
|  |  | Class 4 | 66 | No death observed |  |
|  |  | Class 3 | 748 | All-cause mortality (vs. Ref.)↓ |  |
|  |  | Class 2 | 1,892 | All-cause mortality (vs. Ref.)↓ |  |
|  |  | Class 1 [Least active] | 1,547 | Ref. |  |
| (Yamamoto et al., 2025) | Number of steps, LPA, MVPA, and the ratio of MVPA to total activity | Group-A [low PA] | 407 | Ref. | Logistic regression analysis |
|  |  | Group-B | 81 | No statistically significant difference (vs. Ref.) |  |
|  |  | Group-C | 248 | Metabolic syndrome onset (Ref.)↓ |  |
|  |  | Group-D [high PA] | 58 | No statistically significant difference (vs. Ref.) |  |
| (Albalak et al., 2023) | Mean hourly relative acceleration over a 24-hour period across all measurement days | Average pattern | 27,552 | Ref. | Cox proportional hazards models |
|  |  | Early morning peak | 19,843 | Incident coronary artery disease (vs. Ref.)↓ |  |
|  |  | Late morning peak | 22,648 | Incident coronary artery disease (vs. Ref.)↓, Incident stroke (vs. Ref.)↓, Incident ischemic stroke (vs. Ref.)↓ |  |
|  |  | Evening peak | 16,614 | No statistically significant difference (vs. Ref.) |  |
| (Q. Feng et al., 2025) | Mean hourly relative acceleration over a 24-hour period across all measurement days | Mid-day average pattern | 27,681 | Ref. | Cox proportional hazards models |
|  |  | Early morning peak | 18,614 | No statistically significant difference (vs. Ref.) |  |
|  |  | Late morning peak | 23,299 | Incident type 2 diabetes (vs. Ref.)↓ |  |
|  |  | Early evening peak | 19,845 | No statistically significant difference (vs. Ref.) |  |
| (Bai, Shao, et al., 2024) | Mean hourly acceleration over a 24-hour period across all measurement days | Consistently low | 13,764 | Ref. | Cox proportional hazards models |
|  |  | Single peak | 47,500 | Incident type 2 diabetes (vs. Ref.)↓ |  |
|  |  | Double peak | 18,681 | Incident type 2 diabetes (vs. Ref.)↓ |  |
|  |  | Intense | 19,587 | Incident type 2 diabetes (vs. Ref.)↓ |  |
| (Li et al., 2024) | Mean hourly acceleration over a 24-hour period across all measurement days | PA peak at around 10 a.m. every day | 37,216 | Incident ischemic heart disease (vs. Ref.)↓ | Cox proportional hazards models |
|  |  | Peaks at around 8 a.m. and 6 p.m. daily | 20,045 | Incident ischemic heart disease (vs. Ref.)↓ |  |
|  |  | A relatively uniform distribution of physical intensity throughout the day | 12,666 | Ref. |  |
|  |  | Even daily distribution of PA intensity | 690 | No statistically significant difference (vs. Ref.) |  |
| (Ma et al., 2023) | Mean hourly proportion of MVPA over a 24-hour period | Morning [MVPA] | 642 | Ref. | Linear regression models |
|  |  | Midday [MVPA] | 2,456 | BMI (vs. Ref.)↑, WC (vs. Ref.)↑ |  |
|  |  | Evening [MVPA] | 2,187 | BMI (vs. Ref.)↑, WC (vs. Ref.)↑ |  |
| (Biswas et al., 2023) | Average counts per minute over 10-minute intervals throughout the 24-hour period | Lowest activity | 2,808 | Ref. | Ordinary least squares regression models |
|  |  | Moderate consistent activity | 3,219 | WC (vs. Ref.)↓ |  |
|  |  | Fluctuating moderate activity | 1,194 | WC (vs. Ref.)↓, Systolic BP↓, Cholesterol (vs. Ref.)↓ |  |
|  |  | High daytime activity | 713 | WC (vs. Ref.)↓, Cholesterol (vs. Ref.)↓ |  |
|  |  | Moderate evening activity | 225 | No differences vs. Ref. |  |
|  |  | Highest activity | 750 | WC (vs. Ref.)↓, Systolic BP (vs. Ref.)↓, Diastolic BP (vs. Ref.)↓, Cholesterol (vs. Ref.)↓ |  |
| (Yamamoto et al., 2023) | Number of steps, LPA, MVPA, and the ratio of MVPA to total activity | Group-A [low PA] | 1,047 | Ref. | Logistic regression models |
|  |  | Group-B | 261 | No statistically significant difference (vs. Ref.) |  |
|  |  | Group-C | 418 | Metabolic syndrome (vs. Ref.)↓, Abdominal obesity (vs. Ref.)↓ |  |
|  |  | Group-D [high PA] | 124 | Metabolic syndrome (vs. Ref.)↓, Abdominal obesity (vs. Ref.)↓ |  |
| (Biswas et al., 2022) | Average counts per minute over 10-minute intervals throughout the 24-hour period. | Lowest activity | 2,808 | Ref. | Robust Poisson regression models |
|  |  | Moderate activity | 3,219 | 10-year risk for atherosclerotic CVD event (vs. Ref.)↓ |  |
|  |  | Fluctuations of moderate activity | 1,194 | 10-year risk for atherosclerotic CVD event (vs. Ref.)↓ |  |
|  |  | High daytime activity | 713 | No statistically significant difference (vs. Ref.) |  |
|  |  | Moderate evening activity | 225 | 10-year risk for atherosclerotic CVD event (vs. Ref.)↓ |  |
|  |  | Highest activity | 750 | 10-year risk for atherosclerotic CVD event (vs. Ref.)↓ |  |
| <sup>2</sup>(Leskinen et al., 2023) | Hourly stepping minutes of two consecutive hours | Consistently inactive | 1,301 | BMI (vs. Ref.)↑, Waist (vs. Ref.)↑, 2-hour glucose (vs. Ref.)↑, Total/HDL ratio (vs. Ref.)↑, Triglycerides (vs. Ref.)↑ | Analysis of covariance (ANCOVA) |
|  |  | Consistently low active | 2,498 | BMI (vs. Ref.)↑, Waist (vs. Ref.)↑, 2-hour glucose (vs. Ref.)↑, Total/HDL ratio (vs. Ref.)↑, Triglycerides (vs. Ref.)↑ |  |
|  |  | Active on weekdays | 881 | BMI (vs. Ref.)↑, Waist (vs. Ref.)↑ |  |
|  |  | Early birds | 138 | BMI (vs. Ref.)↑, Waist (vs. Ref.)↑, Systolic BP (vs. Ref.)↑, Diastolic BP (vs. Ref.)↑ |  |
|  |  | Consistently moderately active | 449 | No statistically significant difference (vs. Ref.) |  |
|  |  | Weekend warriors | 453 | BMI (vs. Ref.)↑, Waist (vs. Ref.)↑ |  |
|  |  | Consistently highly active | 352 | Ref. |  |
| (Aqeel et al., 2021) | Summation of hourly activity counts and time stamps | Lowest activity counts from 6:00–23:00 | 783 | Ref. | Linear regression models |
|  |  | High activity counts with a decline after 18:00 | 247 | BMI (vs. Ref.)↓, WC (vs. Ref.)↓ |  |
|  |  | Lower activity counts between 6:00–12:00 | 336 | BMI (vs. Ref.)↓, WC (vs. Ref.)↓, Hemoglobin A1c (vs. Ref.)↓ |  |
|  |  | Higher activity counts between 8:00–11:00 | 633 | BMI (vs. Ref.)↓, WC (vs. Ref.)↓ |  |
| <sup>3</sup>(Niemelä et al., 2019) | Average MET (metabolic equivalent) over 10-minute intervals throughout the 24-hour period | Inactive | 1,881 | 10-year CVD risk (vs. Very active)↑, HDL cholesterol (vs. Very active)↑ | Analysis of variance (ANOVA) |
|  |  | Evening active | 802 | No statistically significant difference (vs. any of the profiles) |  |
|  |  | Moderately active | 1,297 | HDL cholesterol (vs. Very active)↓ |  |
|  |  | Very active | 602 | 10-year CVD risk (vs. Inactive)↓, HDL cholesterol (vs. Inactive)↓, HDL cholesterol (vs. Moderately active)↑ |  |
| (Marschollek, 2016) |  | Cluster 1: High overall physical with regular low-intensity | 92 | BMI (vs. Cluster 3)↓ | Analysis of variance (ANOVA) |
|  | PA intensity, mean duration of PA per 1000 min, and PA regularity | activity and more irregular high-intensity activity |  |  |  |
|  |  | Cluster 2: Regularly and extensively active | 2,230 | C-reactive protein (vs. Cluster 3 and Cluster 4)↓, BMI (vs. Cluster 3 and Cluster 4)↓ |  |
|  |  | Cluster 3: Irregularly and less extensively active | 2,990 | C-reactive protein (vs. Cluster 4)↓, BMI (vs. Cluster 4)↓, C-reactive protein (vs. Cluster 2)↑, BMI (vs. Cluster 1 and Cluster 2)↑ |  |
|  |  | Cluster 4: Inactive | 2,010 | C-reactive protein (vs. Cluster 2 and Cluster 3)↑, BMI (vs. Cluster 1, Cluster 2, and Cluster 3)↑ |  |
| (P. H. Lee et al., 2013) | Hourly average counts per minute over a 24-hour period on weekdays and weekends | Active | 480 | Ref. | Logistic regression models |
|  |  | Less active | 1,234 | Having been diagnosed with a chronic condition (vs. Ref.)↑, Asthma (vs. Ref.)↑ |  |
| (Fukuoka et al., 2018) | Raw minute-level MET for 7 consecutive days | Morning engaged | 65 | Depression symptom (vs. Afternoon)↑; Depression symptom (vs. Unengaged)↓ | Chi-square or analysis of variance (ANOVA) |
|  |  | Afternoon engaged | 48 | Depression symptom (vs. Morning engaged)↓ |  |
|  |  | Unengaged | 102 | Depression symptom (vs. Afternoon and Morning engaged)↑ |  |
| (Nawrin et al., 2024) | Hourly activity counts over a 24-hour period | Morning dominant | 1,964 | Ref. | Logistic regression models |
|  |  | All-day dominant | 1,792 | No apparent differences in health outcomes |  |
|  |  | All-day or morning dominant | 1,855 | No apparent differences in health outcomes |  |
|  |  | Evening dominant | 1,002 | Depressive symptoms↑ |  |
| (Bai, Ning, et al., 2024) | Mean hourly acceleration over a 24-hour period across all measurement days | Persistent low | 14,037 | Ref. | Cox proportional hazards models |
|  |  | Single peak | 46,100 | Incident breast cancer (vs. Ref.)↓, Incident kidney cancer (vs. Ref.)↓, Incident myeloma cancer (vs. Ref.)↓ |  |
|  |  | Double peak | 17,763 | Incident colorectal cancer (vs. Ref.)↓, Incident kidney cancer (vs. Ref.)↓ |  |
|  |  | Vigorous | 18,787 | Incident bladder cancer (vs. Ref.)↓, Incident breast cancer (vs. Ref.)↓, Incident kidney cancer (vs. Ref.)↓, Incident lung cancer (vs. Ref.)↓, Incident myeloma cancer (vs. Ref.)↓, Incident oral cavity and pharynx cancer (vs. Ref.)↓ |  |

Only statistically significant associations observed in the studies are reported. Upward arrow (↑) indicates higher values/incidence in the health outcome relative to the referent group, and downward arrow (↓) lower values/incidence in the health outcome relative to the referent group. <sup>1</sup>: Five models were created based on different input variables into the profiling approach; associations reported here are for the profiles identified on the basis of average activity counts/minute. <sup>2</sup>: The analyses were stratified by type 2 diabetes status; significant associations reported here are those that were seen for no diabetes group <sup>3</sup>: Analysis were stratified by gender; significant differences reported here are those that were seen in both males and females. Abbreviations: Ref. = Referent profile; BMI = Body mass index; BP = Blood pressure; CVD = Cardiovascular diseases; HDL = High-density lipoprotein; LDL = Low-density lipoprotein; LPA= Light-intensity physical activity; MET = Metabolic equivalents; MIMS = Monitor Independent Movement Summary unit; MVPA = Moderate-to-vigorous physical activity; PA = Physical activity; WC = Waist circumference.

All but one (Bai, Zhou, et al., 2024) of the studies creating PA profiles (n = 21), specified the accelerometer-derived descriptors used as input for profile analysis. PA profiling was most commonly conducted using hourly metrics derived from accelerometer outputs, although the specific summary metrics varied across studies. Of the 21 studies, 12 utilized average hourly summary metrics to drive PA profiles (Albalak et al., 2023; Aqeel et al., 2021; Bai, Ning, et al., 2024; Bai, Shao, et al., 2024; Fabrizio et al., 2024; Q. Feng et al., 2025; P. H. Lee et al., 2013; Li et al., 2024; Ma et al., 2023; Nawrin et al., 2024; Shim et al., 2023; Zhang et al., 2024). These 12 studies computed the summary metrics for each hour of the 24-hour day. The summary metrics included hourly average acceleration (Albalak et al., 2023; Bai, Ning, et al., 2024; Bai, Shao, et al., 2024; Fabrizio et al., 2024; Q. Feng et al., 2025; Li et al., 2024), hourly monitor-independent movement summary (MIMS) unit (Shim et al., 2023; Zhang et al., 2024), hourly activity counts (Aqeel et al., 2021; Nawrin et al., 2024), and hourly moderate-to-vigorous PA (MVPA) minutes (Ma et al., 2023). One study used hourly activity counts calculated separately over weekdays and weekend days (P. H. Lee et al., 2013). Four other studies computed summary metrics that were aggregated over different time intervals than on an hourly basis (Biswas et al., 2022, 2023; Leskinen et al., 2023; Niemelä et al., 2019).

### Key profile characteristics and their relationships with health outcomes

Of the 22 studies identifying PA profiles, 14 identified PA profiles that were characterized by having the lowest level of PA relative to other PA profiles in the sample study population. These profiles are depicted in orange in Figure 2. Conversely, of the 22 studies identifying PA profiles, 13 identified PA profiles that were distinguished by having the highest level of PA relative to other identified profiles in the sample study population. These profiles are shown in black in Figure 2.

When a low PA profile emerged, it was generally one of the largest profiles in terms of participant numbers, with few exceptions (Bai, Ning, et al., 2024; Bai, Shao, et al., 2024; Bai, Zhou, et al., 2024). In contrast to low PA profiles, high PA profiles represented a smaller group in terms of participant numbers (see Figure 2). High PA profiles were found to have the most favorable health outcomes, exhibiting reduced risks of all-cause mortality (Bai, Zhou, et al., 2024; Evenson et al., 2017; Shim et al., 2023), incident CVD and CVD mortality (Bai, Zhou, et al., 2024), incident type 2 diabetes (Bai, Shao, et al., 2024), and certain types of cancer (Bai, Ning, et al., 2024), and better markers of cardiometabolic health and obesity (Biswas et al., 2022, 2023; P. H. Lee et al., 2013; Leskinen et al., 2023; Marschollek, 2016; Niemelä et al., 2019; Yamamoto et al., 2023).

Across all studies performing PA profiling, 17 identified at least one profile whose PA peaked at a certain time within a 24-hour cycle. Thirteen of these studies performed with six unique sample populations identified a profile that could be characterized by having a relatively higher PA peak at some time during morning. These profiles are highlighted in green in Figure 2. Profiles of afternoon or evening PA characterized by having a greater proportion of PA in the afternoon or evening emerged in 10 studies performed with six unique sample populations. These profiles are highlighted in yellow in Figure 2. Five studies, conducted across four distinct sample populations, identified profiles characterized by relatively higher physical activity during the day (Biswas et al., 2022, 2023; Q. Feng et al., 2025; Ma et al., 2023; Zhang et al., 2024). These profiles are highlighted in gray in Figure 2.

There were substantial variations among the studies, making it difficult to compare the profiles with respect to the exact timing of peak PA. Those studies identifying morning profiles observed that PA in morning profiles generally peaked *approximately* at some time in between 5:00 and 10:00 (Albalak et al., 2023; Biswas et al., 2022, 2023; Ma et al., 2023; Zhang et al., 2024) or 9:00 and 12:00 (Albalak et al., 2023; Aqeel et al., 2021; Bai, Ning, et al., 2024; Bai, Shao, et al., 2024; Bai, Zhou, et al., 2024; Fukuoka et al., 2018; Leskinen et al., 2023; Li et al., 2024; Nawrin et al., 2024). The PA peak in afternoon and evening profiles *approximately* occurred at some time in between 18:00 and 21:00 (Albalak et al., 2023; Q. Feng et al., 2025; Fukuoka et al., 2018; Ma et al., 2023; Zhang et al., 2024), or showed a steady increase starting sometime between 12:00 and 16:00 and continuing up to nearly midnight (Biswas et al., 2022, 2023; Nawrin et al., 2024; Niemelä et al., 2019). Studies identifying day profiles generally reported relatively higher physical activity from morning until the end of day around 17:00–18:00.

The association of morning, day, afternoon, and evening profiles with health outcomes was somewhat mixed and different, depending on the health outcomes (see Table 2). One study reported that the “Early morning” profile, but not the “Late afternoon,” had a higher mortality risk compared with the “Midday” profile (Zhang et al., 2024). Two studies were performed with the same sample populations and identified the same profile (Albalak et al., 2023; Q. Feng et al., 2025). One of these reported that both “Early morning peak” and “Late morning peak” had a lower incidence of coronary artery disease compared with “Average pattern,” characterized by having average midday PA (Albalak et al., 2023). In the same study (Albalak et al., 2023), “Late morning peak” was also associated with a lower incidence of stroke and ischemic stroke compared with the “Average pattern.” In the other study (Q. Feng et al., 2025), “Early morning peak,” but not “Late morning,” was associated with a lower incidence of type 2 diabetes compared with “Midday average pattern.” Another study reported that “Morning engaged” had higher depression symptoms compared with “Afternoon engaged,” while both “Morning engaged” and “Afternoon engaged” had lower depression symptoms compared with the “Unengaged” profile (Fukuoka et al., 2018).

Three studies identified a profile in the same sample population whose PA peaked during the day at around 9:00–10:00 (Bai, Ning, et al., 2024; Bai, Shao, et al., 2024; Bai, Zhou, et al., 2024). These reported that the single peak profile was associated with lower all-cause mortality, CVD mortality, stroke, and ischemic heart disease incidence compared with the profile with the lowest PA (Bai, Zhou, et al., 2024), a lower incidence of type 2 diabetes compared with the profile with minimal PA (Bai, Shao, et al., 2024), and a lower incidence of breast, kidney, and myeloma cancer incidence compared with the profile with the lowest PA (Bai, Ning, et al., 2024).

One study reported that “High daytime activity,” but not “Moderate evening activity,” was associated with lower waist circumference and cholesterol levels when compared with the lowest activity profile (Biswas et al., 2023). In contrast, another study using the same sample population found no associations for “High daytime activity” relative to the lowest activity profile, while “Moderate evening activity” was linked to a lower composite 10-year CVD score (Biswas et al., 2022). In another study, the “Evening active” profile did not have differences in terms of cardiometabolic health markers or composite 10-year CVD score when compared with all other profiles, including the profile with the lowest activity level (Niemelä et al., 2019). However, one study reported that “Evening (MVPA)” had higher BMI and waist circumference compared with both “Midday (MVPA)” and “Morning (MVPA)” (Ma et al., 2023). Another study identifying an “Evening dominant” profile observed higher depression symptoms for this profile when compared with the “Morning dominant” profile (Nawrin et al., 2024).

### Profiles of sedentary behavior

The overview of profiles identified in four studies creating sedentary behavior profiles is shown in Figure 3. The number of profiles that emerged in these studies ranged from three to five. The input descriptor variables used for profile analysis, the derived sedentary profiles, and their associations with health outcomes in each study are listed in Table 3.

**Figure 3:**
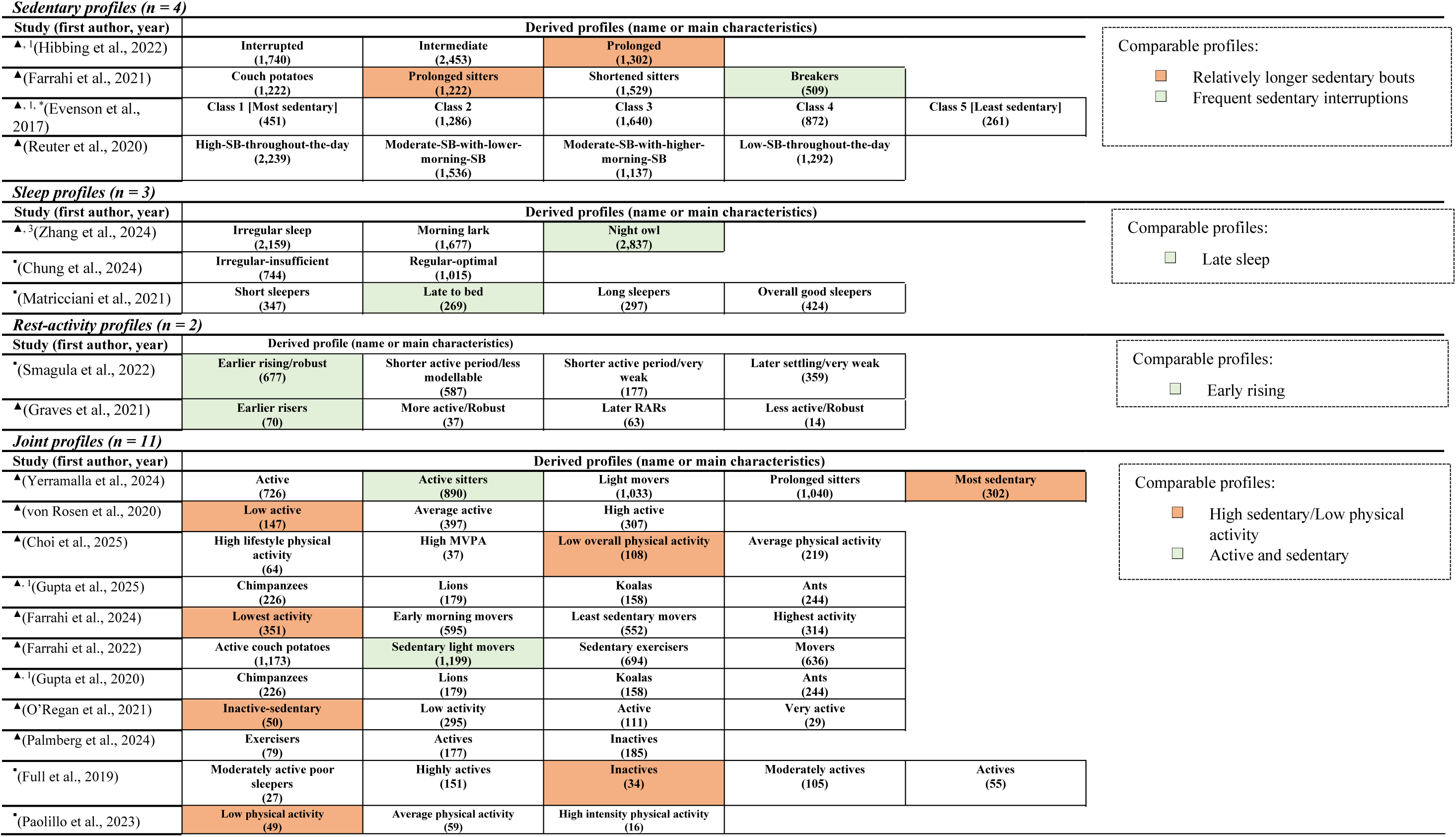
Overview of derived profiles in studied identifying sedentary (n = 4), sleep (n = 2), rest-activity (n = 3), and joint (n = 11) profiles. The numbers shown in () are the number of participants in the profile. The profiles are specified according to the names given to them in the study or a short outline of the main characteristics. To facilitate comparison across studies, profiles with comparable overall characteristics *seen in at least two or more distinct sample populations* are highlighted with distinct colors. Note that the color-coded profiles are not necessarily exactly identical, because of the underlying heterogeneities related to profile analysis and/or accelerometer data cleaning/processing. 1: studies marked with the same number were performed with the same sample population. ▲: High quality study. ▪: Moderate quality study. Individual publication quality scores are shown in Supplementary Table S2. *: Two models were created based on different input variables into the profiling approach; profiles reported here are for the profiles identified on the basis of percentage of sedentary behavior out of total wear time per day. Abbreviations: SB = sedentary behavior; MVPA = Moderate-to-vigorous physical activity.

**Table 3:**
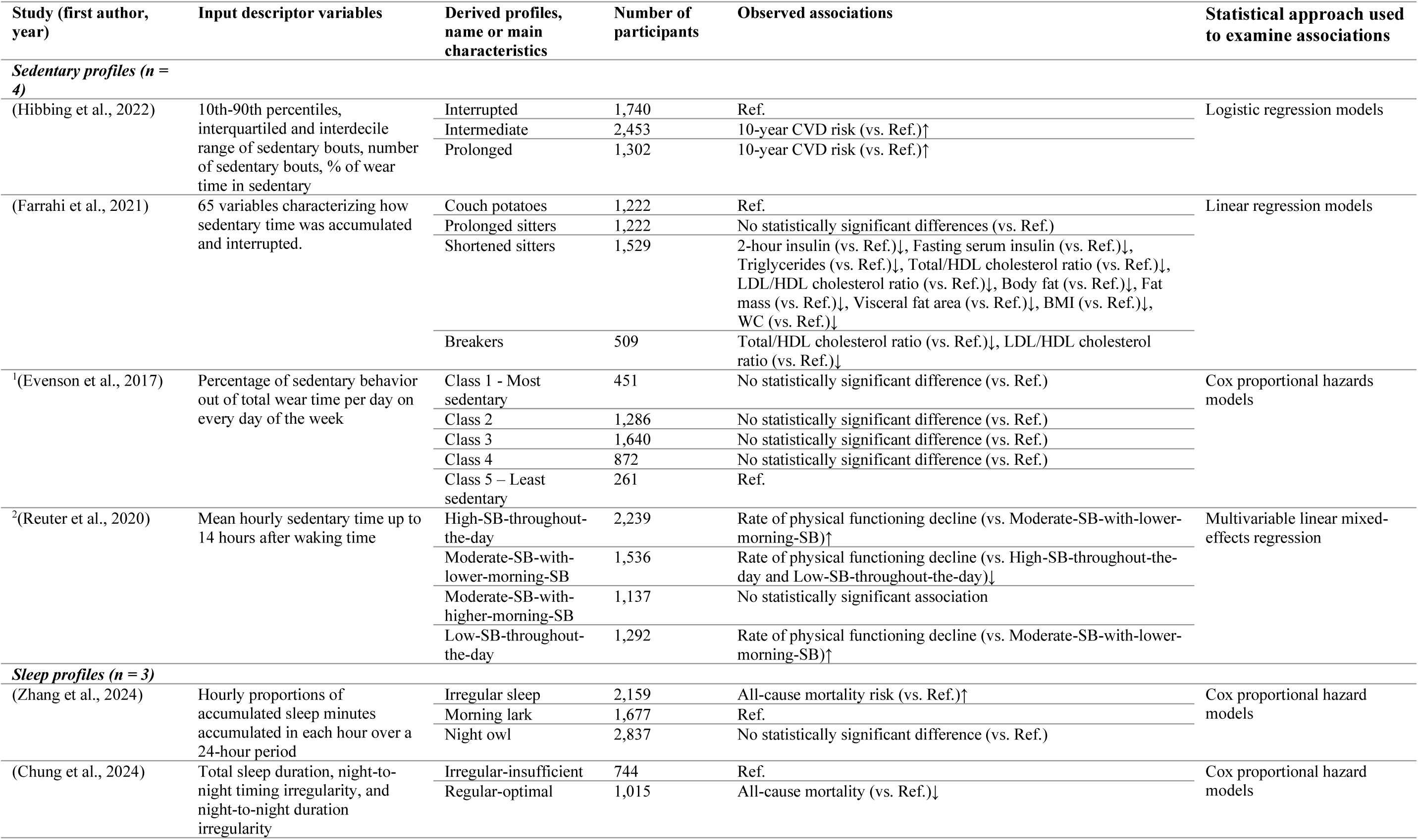
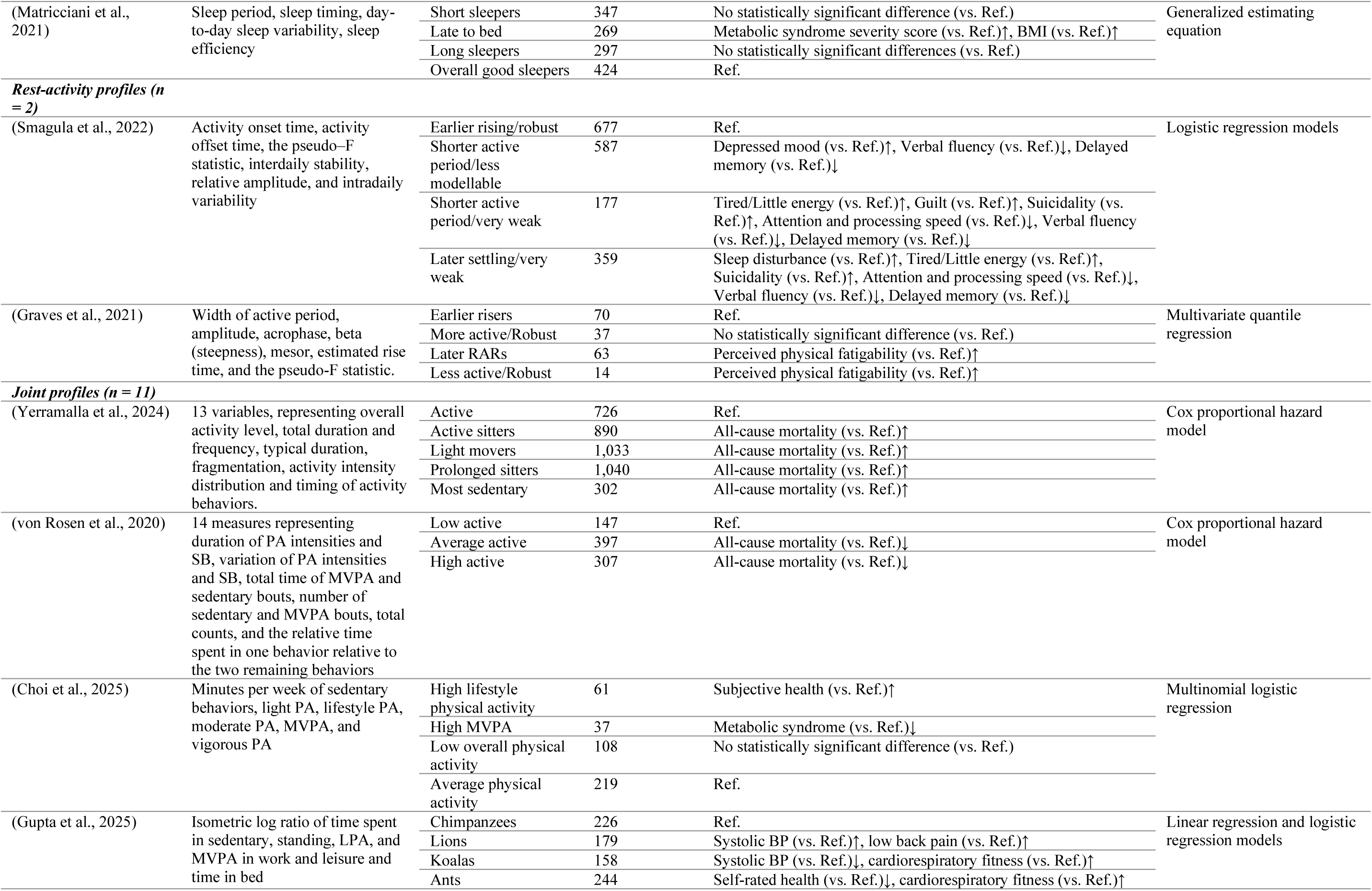
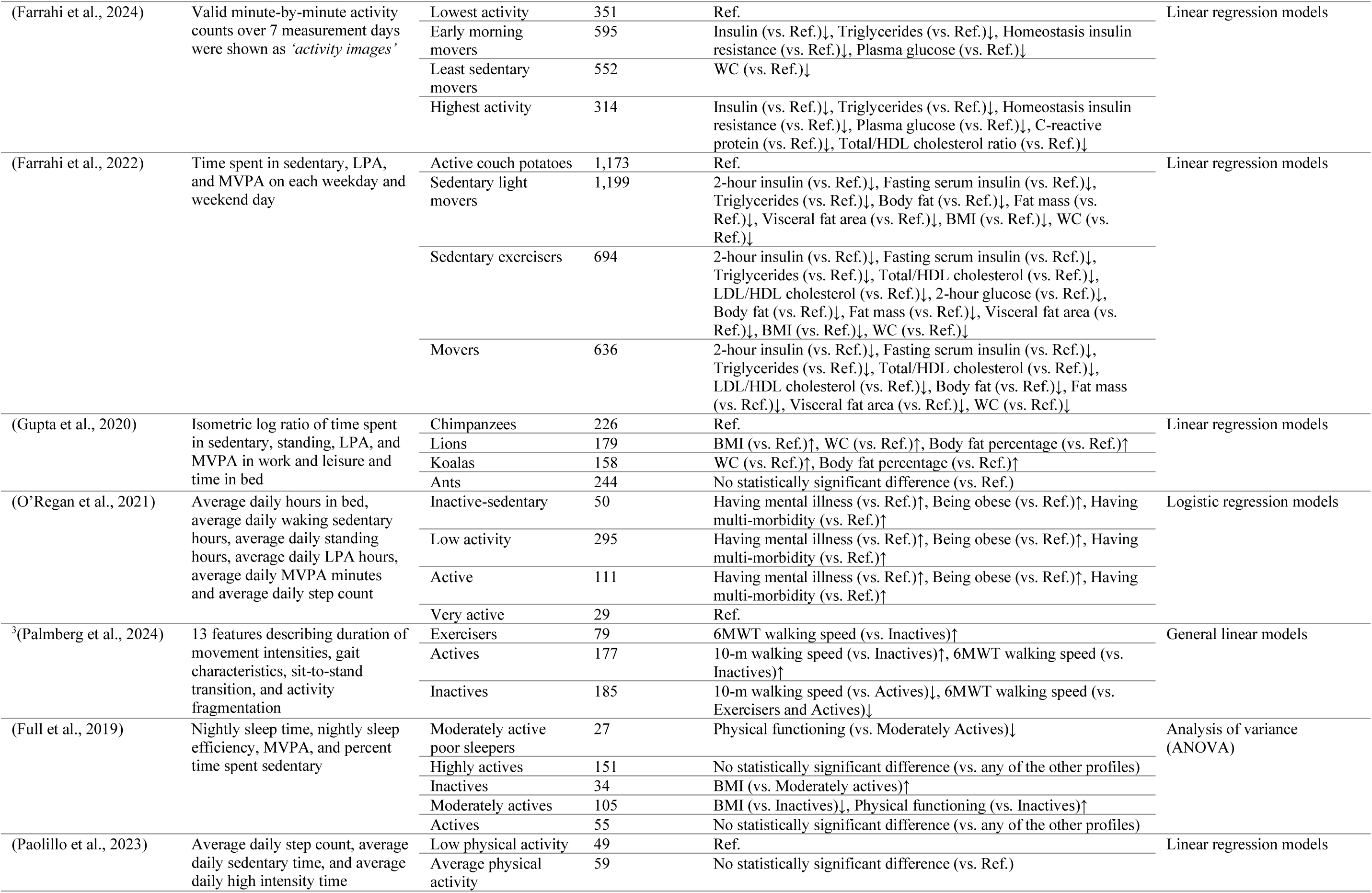
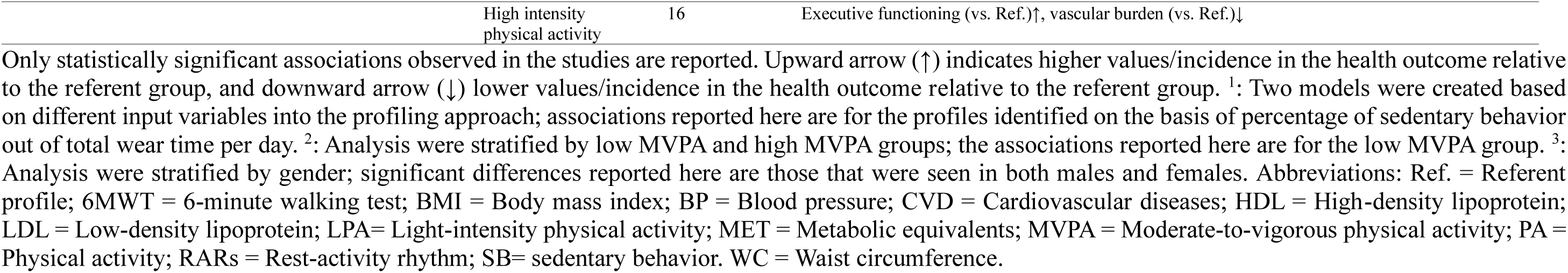
Input descriptor variables, profiles identified, and the statistically significant associations observed among the profiles in studies identifying sedentary behavior, sleep, rest-activity, and joint profiles.

| Study (first author, year) | Input descriptor variables | Derived profiles, name or main characteristics | Number of participants | Observed associations | Statistical approach used to examine associations |
| --- | --- | --- | --- | --- | --- |
| *Sedentary profiles (n = 4)* |  |  |  |  |  |
| (Hibbing et al., 2022) | 10th-90th percentiles, interquartiled and interdecile range of sedentary bouts, number of sedentary bouts, % of wear time in sedentary | Interrupted | 1,740 | Ref. | Logistic regression models |
|  |  | Intermediate | 2,453 | 10-year CVD risk (vs. Ref.)↑ |  |
|  |  | Prolonged | 1,302 | 10-year CVD risk (vs. Ref.)↑ |  |
| (Farrahi et al., 2021) | 65 variables characterizing how sedentary time was accumulated and interrupted. | Couch potatoes | 1,222 | Ref. | Linear regression models |
|  |  | Prolonged sitters | 1,222 | No statistically significant differences (vs. Ref.) |  |
|  |  | Shortened sitters | 1,529 | 2-hour insulin (vs. Ref.)↓, Fasting serum insulin (vs. Ref.)↓, Triglycerides (vs. Ref.)↓, Total/HDL cholesterol ratio (vs. Ref.)↓, LDL/HDL cholesterol ratio (vs. Ref.)↓, Body fat (vs. Ref.)↓, Fat mass (vs. Ref.)↓, Visceral fat area (vs. Ref.)↓, BMI (vs. Ref.)↓, WC (vs. Ref.)↓ |  |
|  |  | Breakers | 509 | Total/HDL cholesterol ratio (vs. Ref.)↓, LDL/HDL cholesterol ratio (vs. Ref.)↓ |  |
| <sup>1</sup>(Evenson et al., 2017) | Percentage of sedentary behavior out of total wear time per day on every day of the week | Class 1 - Most sedentary | 451 | No statistically significant difference (vs. Ref.) | Cox proportional hazards models |
|  |  | Class 2 | 1,286 | No statistically significant difference (vs. Ref.) |  |
|  |  | Class 3 | 1,640 | No statistically significant difference (vs. Ref.) |  |
|  |  | Class 4 | 872 | No statistically significant difference (vs. Ref.) |  |
|  |  | Class 5 – Least sedentary | 261 | Ref. |  |
| <sup>2</sup>(Reuter et al., 2020) | Mean hourly sedentary time up to 14 hours after waking time | High-SB-throughout-the-day | 2,239 | Rate of physical functioning decline (vs. Moderate-SB-with-lower-morning-SB)↑ | Multivariable linear mixed-effects regression |
|  |  | Moderate-SB-with-lower-morning-SB | 1,536 | Rate of physical functioning decline (vs. High-SB-throughout-the-day and Low-SB-throughout-the-day)↓ |  |
|  |  | Moderate-SB-with-higher-morning-SB | 1,137 | No statistically significant association |  |
|  |  | Low-SB-throughout-the-day | 1,292 | Rate of physical functioning decline (vs. Moderate-SB-with-lower-morning-SB)↑ |  |
| *Sleep profiles (n = 3)* |  |  |  |  |  |
| (Zhang et al., 2024) | Hourly proportions of accumulated sleep minutes accumulated in each hour over a 24-hour period | Irregular sleep | 2,159 | All-cause mortality risk (vs. Ref.)↑ | Cox proportional hazard models |
|  |  | Morning lark | 1,677 | Ref. |  |
|  |  | Night owl | 2,837 | No statistically significant difference (vs. Ref.) |  |
| (Chung et al., 2024) | Total sleep duration, night-to-night timing irregularity, and night-to-night duration irregularity | Irregular-insufficient | 744 | Ref. | Cox proportional hazard models |
|  |  | Regular-optimal | 1,015 | All-cause mortality (vs. Ref.)↓ |  |
| (Matricciani et al., 2021) | Sleep period, sleep timing, day-to-day sleep variability, sleep efficiency | Short sleepers | 347 | No statistically significant difference (vs. Ref.) | Generalized estimating equation |
|  |  | Late to bed | 269 | Metabolic syndrome severity score (vs. Ref.)↑, BMI (vs. Ref.)↑ |  |
|  |  | Long sleepers | 297 | No statistically significant differences (vs. Ref.) |  |
|  |  | Overall good sleepers | 424 | Ref. |  |
| **Rest-activity profiles (n = 2)** |  |  |  |  |  |
| (Smagula et al., 2022) | Activity onset time, activity offset time, the pseudo-F statistic, interdaily stability, relative amplitude, and intradaily variability | Earlier rising/robust | 677 | Ref. | Logistic regression models |
|  |  | Shorter active period/less modellable | 587 | Depressed mood (vs. Ref.)↑, Verbal fluency (vs. Ref.)↓, Delayed memory (vs. Ref.)↓ |  |
|  |  | Shorter active period/very weak | 177 | Tired/Little energy (vs. Ref.)↑, Guilt (vs. Ref.)↑, Suicidality (vs. Ref.)↑, Attention and processing speed (vs. Ref.)↓, Verbal fluency (vs. Ref.)↓, Delayed memory (vs. Ref.)↓ |  |
|  |  | Later settling/very weak | 359 | Sleep disturbance (vs. Ref.)↑, Tired/Little energy (vs. Ref.)↑, Suicidality (vs. Ref.)↑, Attention and processing speed (vs. Ref.)↓, Verbal fluency (vs. Ref.)↓, Delayed memory (vs. Ref.)↓ |  |
| (Graves et al., 2021) | Width of active period, amplitude, acrophase, beta (steepness), mesor, estimated rise time, and the pseudo-F statistic. | Earlier risers | 70 | Ref. | Multivariate quantile regression |
|  |  | More active/Robust | 37 | No statistically significant difference (vs. Ref.) |  |
|  |  | Later RARs | 63 | Perceived physical fatigability (vs. Ref.)↑ |  |
|  |  | Less active/Robust | 14 | Perceived physical fatigability (vs. Ref.)↑ |  |
| **Joint profiles (n = 11)** |  |  |  |  |  |
| (Yerramalla et al., 2024) | 13 variables, representing overall activity level, total duration and frequency, typical duration, fragmentation, activity intensity distribution and timing of activity behaviors. | Active | 726 | Ref. | Cox proportional hazard model |
|  |  | Active sitters | 890 | All-cause mortality (vs. Ref.)↑ |  |
|  |  | Light movers | 1,033 | All-cause mortality (vs. Ref.)↑ |  |
|  |  | Prolonged sitters | 1,040 | All-cause mortality (vs. Ref.)↑ |  |
|  |  | Most sedentary | 302 | All-cause mortality (vs. Ref.)↑ |  |
| (von Rosen et al., 2020) | 14 measures representing duration of PA intensities and SB, variation of PA intensities and SB, total time of MVPA and sedentary bouts, number of sedentary and MVPA bouts, total counts, and the relative time spent in one behavior relative to the two remaining behaviors | Low active | 147 | Ref. | Cox proportional hazard model |
|  |  | Average active | 397 | All-cause mortality (vs. Ref.)↓ |  |
|  |  | High active | 307 | All-cause mortality (vs. Ref.)↓ |  |
| (Choi et al., 2025) | Minutes per week of sedentary behaviors, light PA, lifestyle PA, moderate PA, MVPA, and vigorous PA | High lifestyle physical activity | 61 | Subjective health (vs. Ref.)↑ | Multinomial logistic regression |
|  |  | High MVPA | 37 | Metabolic syndrome (vs. Ref.)↓ |  |
|  |  | Low overall physical activity | 108 | No statistically significant difference (vs. Ref.) |  |
|  |  | Average physical activity | 219 | Ref. |  |
| (Gupta et al., 2025) | Isometric log ratio of time spent in sedentary, standing, LPA, and MVPA in work and leisure and time in bed | Chimpanzees | 226 | Ref. | Linear regression and logistic regression models |
|  |  | Lions | 179 | Systolic BP (vs. Ref.)↑, low back pain (vs. Ref.)↑ |  |
|  |  | Koalas | 158 | Systolic BP (vs. Ref.)↓, cardiorespiratory fitness (vs. Ref.)↑ |  |
|  |  | Ants | 244 | Self-rated health (vs. Ref.)↓, cardiorespiratory fitness (vs. Ref.)↑ |  |
| (Farrahi et al., 2024) | Valid minute-by-minute activity counts over 7 measurement days were shown as 'activity images' | Lowest activity | 351 | Ref. | Linear regression models |
|  |  | Early morning movers | 595 | Insulin (vs. Ref.)↓, Triglycerides (vs. Ref.)↓, Homeostasis insulin resistance (vs. Ref.)↓, Plasma glucose (vs. Ref.)↓ |  |
|  |  | Least sedentary movers | 552 | WC (vs. Ref.)↓ |  |
|  |  | Highest activity | 314 | Insulin (vs. Ref.)↓, Triglycerides (vs. Ref.)↓, Homeostasis insulin resistance (vs. Ref.)↓, Plasma glucose (vs. Ref.)↓, C-reactive protein (vs. Ref.)↓, Total/HDL cholesterol ratio (vs. Ref.)↓ |  |
| (Farrahi et al., 2022) | Time spent in sedentary, LPA, and MVPA on each weekday and weekend day | Active couch potatoes | 1,173 | Ref. | Linear regression models |
|  |  | Sedentary light movers | 1,199 | 2-hour insulin (vs. Ref.)↓, Fasting serum insulin (vs. Ref.)↓, Triglycerides (vs. Ref.)↓, Body fat (vs. Ref.)↓, Fat mass (vs. Ref.)↓, Visceral fat area (vs. Ref.)↓, BMI (vs. Ref.)↓, WC (vs. Ref.)↓ |  |
|  |  | Sedentary exercisers | 694 | 2-hour insulin (vs. Ref.)↓, Fasting serum insulin (vs. Ref.)↓, Triglycerides (vs. Ref.)↓, Total/HDL cholesterol (vs. Ref.)↓, LDL/HDL cholesterol (vs. Ref.)↓, 2-hour glucose (vs. Ref.)↓, Body fat (vs. Ref.)↓, Fat mass (vs. Ref.)↓, Visceral fat area (vs. Ref.)↓, BMI (vs. Ref.)↓, WC (vs. Ref.)↓ |  |
|  |  | Movers | 636 | 2-hour insulin (vs. Ref.)↓, Fasting serum insulin (vs. Ref.)↓, Triglycerides (vs. Ref.)↓, Total/HDL cholesterol (vs. Ref.)↓, LDL/HDL cholesterol (vs. Ref.)↓, Body fat (vs. Ref.)↓, Fat mass (vs. Ref.)↓, Visceral fat area (vs. Ref.)↓, WC (vs. Ref.)↓ |  |
| (Gupta et al., 2020) | Isometric log ratio of time spent in sedentary, standing, LPA, and MVPA in work and leisure and time in bed | Chimpanzees | 226 | Ref. | Linear regression models |
|  |  | Lions | 179 | BMI (vs. Ref.)↑, WC (vs. Ref.)↑, Body fat percentage (vs. Ref.)↑ |  |
|  |  | Koalas | 158 | WC (vs. Ref.)↑, Body fat percentage (vs. Ref.)↑ |  |
|  |  | Ants | 244 | No statistically significant difference (vs. Ref.) |  |
| (O'Regan et al., 2021) | Average daily hours in bed, average daily waking sedentary hours, average daily standing hours, average daily LPA hours, average daily MVPA minutes and average daily step count | Inactive-sedentary | 50 | Having mental illness (vs. Ref.)↑, Being obese (vs. Ref.)↑, Having multi-morbidity (vs. Ref.)↑ | Logistic regression models |
|  |  | Low activity | 295 | Having mental illness (vs. Ref.)↑, Being obese (vs. Ref.)↑, Having multi-morbidity (vs. Ref.)↑ |  |
|  |  | Active | 111 | Having mental illness (vs. Ref.)↑, Being obese (vs. Ref.)↑, Having multi-morbidity (vs. Ref.)↑ |  |
|  |  | Very active | 29 | Ref. |  |
| <sup>3</sup>(Palmberg et al., 2024) | 13 features describing duration of movement intensities, gait characteristics, sit-to-stand transition, and activity fragmentation | Exercisers | 79 | 6MWT walking speed (vs. Inactives)↑ | General linear models |
|  |  | Actives | 177 | 10-m walking speed (vs. Inactives)↑, 6MWT walking speed (vs. Inactives)↑ |  |
|  |  | Inactives | 185 | 10-m walking speed (vs. Actives)↓, 6MWT walking speed (vs. Exercisers and Actives)↓ |  |
| (Full et al., 2019) | Nightly sleep time, nightly sleep efficiency, MVPA, and percent time spent sedentary | Moderately active poor sleepers | 27 | Physical functioning (vs. Moderately Actives)↓ | Analysis of variance (ANOVA) |
|  |  | Highly actives | 151 | No statistically significant difference (vs. any of the other profiles) |  |
|  |  | Inactives | 34 | BMI (vs. Moderately actives)↑ |  |
|  |  | Moderately actives | 105 | BMI (vs. Inactives)↓, Physical functioning (vs. Inactives)↑ |  |
|  |  | Actives | 55 | No statistically significant difference (vs. any of the other profiles) |  |
| (Paolillo et al., 2023) | Average daily step count, average daily sedentary time, and average daily high intensity time | Low physical activity | 49 | Ref. | Linear regression models |
|  |  | Average physical activity | 59 | No statistically significant difference (vs. Ref.) |  |

|  |  |  |
| --- | --- | --- |
| High intensity<br>physical activity | 16 | Executive functioning (vs. Ref.)↑, vascular burden (vs. Ref.)↓ |

Only statistically significant associations observed in the studies are reported. Upward arrow (↑) indicates higher values/incidence in the health outcome relative to the referent group, and downward arrow (↓) lower values/incidence in the health outcome relative to the referent group. <sup>1</sup>: Two models were created based on different input variables into the profiling approach; associations reported here are for the profiles identified on the basis of percentage of sedentary behavior out of total wear time per day. <sup>2</sup>: Analysis were stratified by low MVPA and high MVPA groups; the associations reported here are for the low MVPA group. <sup>3</sup>: Analysis were stratified by gender; significant differences reported here are those that were seen in both males and females. Abbreviations: Ref. = Referent profile; 6MWT = 6-minute walking test; BMI = Body mass index; BP = Blood pressure; CVD = Cardiovascular diseases; HDL = High-density lipoprotein; LDL = Low-density lipoprotein; LPA= Light-intensity physical activity; MET = Metabolic equivalents; MVPA = Moderate-to-vigorous physical activity; PA = Physical activity; RARs = Rest-activity rhythm; SB= sedentary behavior. WC = Waist circumference.

In one study (Evenson et al., 2017), the daily percentage of total sedentary time from total accelerometer wear time was used to create and identify sedentary behavior profiles. Two studies created sedentary behavior profiles using several variables describing how sedentary time was accumulated and interrupted (Farrahi et al., 2021; Hibbing et al., 2022), but the descriptor variables used in these studies differed. In the other study, sedentary behavior profiles were created using mean hourly sedentary time computed for up to 14 hours after waking (Reuter et al., 2020). This approach allowed the assessment of diurnal patterns of sedentary time.

### Key profile characteristics and their relationships with health outcomes

Of the four studies creating sedentary profiles, two identified sedentary behavior profiles in distinct sample populations based on how sedentary time was accumulated and interrupted (Farrahi et al., 2021; Hibbing et al., 2022). Both studies identified profiles characterized by having more frequent interruptions in sedentary time compared with other emerging profiles. These profiles, labeled as “Interrupted” (Hibbing et al., 2022) and “Breakers” (Farrahi et al., 2021) are highlighted in green in Figure 3 (part Sedentary profiles). In contrast, both studies also identified profiles characterized by prolonged periods of uninterrupted sedentary time, labeled as “Prolonged” (Hibbing et al., 2022) and “Prolonged sitters” (Farrahi et al., 2021). These profiles are highlighted in orange in Figure 3 (part Sedentary profiles). A comparable threshold for defining prolonged sedentary bouts emerged in both studies (i.e., >20 min and >15–30 min). One of these two studies identified two additional unique profiles: “Shortened sitters,” characterized by sedentary time accumulated in relatively short bouts of 15–30 minutes, and “Couch potatoes,” distinguished by the highest overall levels of sedentary time among all identified profiles (Farrahi et al., 2021).

As shown in Table 3, the “Prolonged” profile was associated with a higher 10-year CVD risk compared with the “Interrupted” profile (Hibbing et al., 2022). In the other study (Farrahi et al., 2021), no statistically significant differences in cardiometabolic health markers were observed between the “Couch potatoes” and “Prolonged sitters.” However, both the “Shortened sitters” and “Breakers” demonstrated better cardiometabolic health compared with the “Couch potatoes” (Farrahi et al., 2021). One study identified sedentary behavior profiles based on sedentary time as a proportion of total daily wear time across all days of the week and found no association between these profiles and mortality risk (Evenson et al., 2017).

### Profiles of sleep

The overview of profiles identified in three studies creating sleep profiles is shown in Figure 3. The input descriptor variables used for profile analysis, the derived sleep profiles, and their associations with health outcomes in each study are listed in Table 3. The number of profiles that emerged in these three data-driven sleep profiling studies was two (Chung et al., 2024), three (Zhang et al., 2024) and four (Matricciani et al., 2021). One of these studies utilized hourly proportions of accumulated sleep minutes within each hour over a 24-hour period as descriptor variables for profile analysis (Zhang et al., 2024). Although not identical, the other two studies employed descriptor metrics related to sleep duration, timing, and variability (Chung et al., 2024; Matricciani et al., 2021).

### Key profile characteristics and their relationships with health outcomes

The three studies identifying sleep profiles were conducted with distinct sample populations (Chung et al., 2024; Matricciani et al., 2021; Zhang et al., 2024). Among these, a profile characterized by late bedtime was identified in two studies, as illustrated in green in Figure 3 (part Sleep profile). In one of these two studies, the profiles identified were “Irregular sleep”, “Morning lark”, and “Night owl” (Zhang et al., 2024). “Irregular sleep” was characterized by varying times of sleep onset and awakening throughout the day, “Morning lark” by sleeping early at night and waking early in the morning, and “Night owl” by going to bed late and waking up late (Zhang et al., 2024). “Irregular sleep” was associated with a higher mortality risk compared with the “Morning lark.” In the other study (Matricciani et al., 2021), the identified profiles were “Overall good sleepers,” “Late to bed,” “Long sleepers”, and “Short sleepers.” Among these profiles (Matricciani et al., 2021), the “Late to bed” profile was associated with higher BMI and metabolic syndrome scores compared with the “Overall good sleepers” profile (see Table 3).

### Profiles of rest-activity rhythmicity

The overview of profiles identified in the two studies that created rest-activity rhythmicity profiles is shown in Figure 3. The input descriptor variables used for profile analysis, the derived rest-activity rhythmicity profiles, and their associations with health outcomes in each study are listed in Table 3. Four profiles emerged in both studies that created and identified profiles of rest-activity rhythmicity (Graves et al., 2021; Smagula et al., 2022). The rest-activity rhythm descriptors used as input for profile analysis in both studies largely overlapped and were derived from an extended cosine model (Graves et al., 2021; Smagula et al., 2022).

### Key profile characteristics and their relationships with health outcomes

The two studies identified rest-activity profiles in distinct populations (Graves et al., 2021; Smagula et al., 2022). “Early rising” profile, highlighted in green in Figure 3 (part Rest-activity rhythmicity), was identified in both studies. In one of the studies (Smagula et al., 2022), “Shorter active period/less modelable” and “Shorter active period/very weak,” characterized by starting their daily activities later and settling down earlier, with a relatively less rhythmic 24-hour rest-activity pattern, exhibited higher depression symptoms, compared with “Earlier rising/robust,” which was characterized by an estimated activity onset before 7 AM and a more rhythmic 24-hour rest-activity pattern (Smagula et al., 2022). In the other study (Graves et al., 2021), “Later rest activity rhythms (RARs),” characterized by having a later activity onset, had higher perceived fatiguability compared with “Earlier risers,” characterized by a relatively earlier activity onset. The “Less activity/robust” profile, characterized by having relatively a more rhythmic rest-activity pattern, exhibited higher perceived fatiguability compared with “Earlier risers” (Graves et al., 2021).

### Joint profiles of physical activity, sedentary behavior, and/or sleep

The overview of profiles identified in 11 studies creating joint profiles is shown in Figure 3. The number of profiles that emerged in these studies ranged from three to five. The input descriptor variables used for profile analysis, the derived joint profiles, and their associations with health outcomes in each study are listed in Table 3. Although durations of physical activity and sedentary behavior were commonly used as descriptor variables, there was overall heterogeneity in the descriptors used as input for identifying and creating joint profiles (see Table 3).

Of the 11 studies creating joint profiles, seven incorporated a combination of accelerometer-derived PA and sedentary behavior descriptors for profile analyses (Choi et al., 2025; Farrahi et al., 2022; Gupta et al., 2025; Palmberg et al., 2024; Paolillo et al., 2023; von Rosen et al., 2020; Yerramalla et al., 2024) and three incorporated a combination of accelerometer-derived PA, sedentary behavior, and sleep descriptors (Full et al., 2019; Gupta et al., 2020; O’Regan et al., 2021). One study used transformed accelerometer outputs into what was named *activity images* and used the activity images for profile analysis (Farrahi et al., 2024).

### Key profile characteristics and their relationships with health outcomes

Due to the heterogeneity and variety of input descriptor variables in profile analysis, comparing joint profiles across the included studies in terms of similarities and differences was more challenging than comparing other types of profiles. Most commonly, profiles distinguished by low levels of PA combined with high levels of sedentary time emerged among the studies identifying joint profiles (n = 7) (Choi et al., 2025; Farrahi et al., 2024; Full et al., 2019; O’Regan et al., 2021; Paolillo et al., 2023; von Rosen et al., 2020; Yerramalla et al., 2024). These profiles are marked in orange in Figure 3 (part Joint profiles). Compared to profiles with low levels of PA combined with high levels of sedentary time, relatively more active profiles having lower levels of sedentary time and higher levels of PA, demonstrated reduced risks of mortality (von Rosen et al., 2020; Yerramalla et al., 2024), lower amount of chronic illness and multimorbidity (O’Regan et al., 2021), better cardiometabolic health and obesity indicators (Farrahi et al., 2024; Full et al., 2019), and better cognitive health (Paolillo et al., 2023) and physical capacity outcomes (Paolillo et al., 2023).

Profiles most distinguished from other profiles by having high sedentary time combined with having sufficient PA were seen in two studies conducted with distinct sample populations (Farrahi et al., 2022; Yerramalla et al., 2024). These profiles are marked in green in Figure 3 (part Joint profiles). As shown in Table 3, in one of these studies, the “Active sitters,” most distinguished by having sufficient MVPA but also high sedentary time, were shown to have a higher mortality risk compared with the “Active” profile, characterized by having both lower sedentary time and higher MVPA (Yerramalla et al., 2024). The other study identified an “Active couch potato” profile, most distinguished by having sufficient MVPA coupled with high sedentary time. Compared with this profile, “Sedentary light movers” who engaged in sufficient PA but had relatively less sedentary time and more light-intensity PA were found to have better cardiometabolic health (Farrahi et al., 2022).

## DISCUSSION

The present study systematically identified and reviewed 40 studies that utilized data-driven population segmentation analysis to drive multidimensional physical behavior profiles from wearable accelerometry data in a sample of adult participants. Machine learning K-means clustering algorithm and latent profile analysis were the most commonly employed methods for profiling of physical behavior from accelerometer data. The review of derived profiles from accelerometer-monitored physical behaviors revealed several hypothesis-generating, preliminary evidence about adult physical behavior profiles. Multiple studies have identified data-driven profiles distinguished by the timing of physical activity across the 24-hour day. Some profiles suggest that adults can be physically active while still spending substantial time in sedentary behaviors. In addition to total sedentary time, the patterns of accumulation and interruption of sedentary behavior may also be associated with health risks and outcomes.

Data-driven population segmentation analysis are widely used in a number of different research fields (Woo et al., 2024). Compared with variable-centered approaches (Migueles et al., 2022), one of the main advantage of algorithms used for cluster analysis and population segmentations is that they can naturally accommodate a large number of input variables (Biswas et al., 2022; Farrahi et al., 2021; Kebede et al., 2024; Liu et al., 2023; Niemelä et al., 2019; Weller et al., 2020). In the domains related to health and medicine, data-driven population segmentation analysis have been predominantly employed for examining how different risk factors in multidimensional data may cluster together (Hafkamp et al., 9900; Kang et al., 2014; Liu et al., 2023; Woo et al., 2024). Consistent with previous research in other domains (Hafkamp et al., 9900; Kang et al., 2014; Liu et al., 2023; Woo et al., 2024), our study highlights the utility of data-driven profiling of accelerometer-measured physical behaviors for addressing questions about how different components and dimensions of physical behaviors cluster together, and analyzing whether and how the relative similarities and dissimilarities.

In general, studies used a wide variety of accelerometer-derived descriptors to characterize physical behavior profiles, making direct comparison of profiles difficult. Still, the profiles identified through data-driven methods generally aligned with the well-established evidence observed in studies employing variable-centered approaches. For example, profiles distinguished by relatively a low PA combined with high sedentary time frequently emerged in studies examining how different components of physical behaviors cluster together (Choi et al., 2025; Farrahi et al., 2024; Full et al., 2019; O’Regan et al., 2021; Paolillo et al., 2023; von Rosen et al., 2020; Yerramalla et al., 2024). Consistent with previous findings (Ekelund et al., 2016; Stamatakis et al., 2019), profiles most distinguished by a combination of low PA and high sedentary time were consistently found to be linked with increased risks of CVD and all-cause mortality and adversely to several other risk indicators (Choi et al., 2025; Farrahi et al., 2024; Full et al., 2019; O’Regan et al., 2021; Paolillo et al., 2023; von Rosen et al., 2020; Yerramalla et al., 2024).

Joint profiles identified in data-driven studies suggest that a segment of the adult population may exhibit a behavioral pattern characterized by being physically active while simultaneously engaging in significant amounts of sedentary behavior. Current robust evidence indicates that although prolonged sedentary behavior is associated with increased risks of various chronic conditions and mortality (Biswas et al., 2015), high levels of PA can mitigate some of the health risks linked to excessive sedentary time (Ekelund et al., 2016). In line with our findings, some population-based studies have reported a high prevalence of people meeting PA recommendations while also accumulating high average sedentary time (Collings et al., 2022; Moreno-Llamas et al., 2022). Although further research is needed to better understand how socioeconomic factors may be related to this physical behavior profile, *active yet sedentary* profile appears to be common among adults in high-income and European countries (Collings et al., 2022; Moreno-Llamas et al., 2022). Currently, it remains unclear whether the level of PA among these individuals is sufficient to offset the health risks posed by their sedentary behavior. The findings of this review highlight that *active yet sedentary* profile may be present among adults, underscoring the need for further research to better understand its prevalence and health implications in adults.

Several studies identifying PA profiles found a profile in which PA peaked at sometime during morning, day, afternoon, or evening (Albalak et al., 2023; Aqeel et al., 2021; Bai, Ning, et al., 2024; Bai, Shao, et al., 2024; Bai, Zhou, et al., 2024; Biswas et al., 2022, 2023; Fabrizio et al., 2024; Q. Feng et al., 2025; Fukuoka et al., 2018; Leskinen et al., 2023; Li et al., 2024; Ma et al., 2023; Nawrin et al., 2024; Niemelä et al., 2019; Shim et al., 2023; Zhang et al., 2024). The health implications of these profiles were mixed in terms of whether PA during a certain time of the 24-hour period is more beneficial than at other times. Currently, whether PA at one time of day provides greater health benefits than at another is still not fully understood. Although the optimal time window for PA may be outcome-specific (H. Feng et al., 2023; Keiser et al., 2024), findings from data-driven studies overall support the hypothesis that the timing of PA may influence its health benefits.

Low PA profiles most distinguished by having the lowest levels of PA, were repeatedly identified across studies performing physical activity profiling. These profiles emerged across several sample populations from various countries and often represented one of the largest participant groups among the identified profiles. The health risks associated with insufficient PA are well-documented and extensively studied previously (I.-M. Lee et al., 2012). Current estimates based on self-reported PA indicate that nearly one-third of adults worldwide do not meet the recommended levels of PA (Strain et al., 2024). In line with existing literature (I.-M. Lee et al., 2012), data-driven PA profiles derived from multidimensional accelerometer data consistently indicate that low PA is associated with increased health risks.

Two studies created sedentary profiles based on how sedentary time is accumulated and interrupted, both identifying a profile distinguished by spending sedentary time in relatively longer bouts lasting 15–30 min (Farrahi et al., 2021; Hibbing et al., 2022). Currently, it is well-established that excessive sedentary time, particularly when accumulated in uninterrupted bouts, poses a health risk (Biswas et al., 2015). However, it remains unclear at what point sedentary behavior begins to negatively impact health outcomes (Chastin, Egerton, et al., 2015). Sedentary profiles derived using data-driven techniques provide preliminary, hypothesis-generating evidence suggesting that limiting sedentary time to bouts of approximately 15–30 min may be linked to better cardiometabolic health (Farrahi et al., 2021; Hibbing et al., 2022). This data-driven hypothesis warrants further research to explore whether similar profiles can be observed across different populations and how limiting sedentary bouts to specific durations is linked to other health outcomes.

Only a few studies have created and identified rest-activity rhythmicity or sleep profiles. Due to the limited number of studies and the heterogeneity observed among them, synthesizing evidence regarding the profiles derived from these studies was challenging. In general, there has been a tendency to apply data-driven population segmentation analysis primarily to accelerometer-determined physical behaviors during waking hours. This may be partly because earlier wearable accelerometers were traditionally used to monitor physical behaviors only during waking hours, with relatively recent studies adopting a 24-hour wear protocol (Evenson et al., 2022; Giurgiu et al., 2025). Although evidence remains inconclusive, early findings from data-driven profiles of sleep suggest that patterns of bedtime and wake-up time may be linked to mortality risk and cardiometabolic health (Matricciani et al., 2021; Zhang et al., 2024). Studies identifying rest-activity profiles also provided some support for this hypothesis indicating that earlier activity onset, compared with later activity onset, may be favorably linked with depressive symptoms (Smagula et al., 2022) and perceived fatiguability (Graves et al., 2021). However, further research is warranted to clarify the potential bidirectional relationships between rest-activity and sleep profiles and health outcomes.

This review study has several notable strengths. An extensive literature search was conducted and both prospective cohort and cross-sectional studies were included. While a previously conducted systematic scoping review has described the application of latent profile analysis to accelerometry data based on a total of 12 included studies (Brusco et al., 2017), our study is significantly more inclusive. It is the first to systematically review studies that identify physical behavior profiles from wearable accelerometry data in adult participants using data-driven population segmentation analysis. Multiple data-driven hypotheses were identified from the review of physical behavior profiles and summarized their relationships with various health outcomes.

This study is not without limitations. In our review, title and abstract screening was conducted by a single reviewer, which may have increased the risk of missed eligible studies. However, single-reviewer screening is often used to reduce the substantial resources required for systematic reviews and has been suggested to be sufficient when performed by a senior expert (Waffenschmidt et al., 2019). In general, profiles derived from accelerometer-measured physical behaviors in different sample populations seem to be dependent on the descriptor variables used as input for profile analysis. The decisions made for cleaning and preparing accelerometer data, as well as measurement specification and accelerometer wear location, could potentially have influenced the final derived profiles within the included studies (Migueles et al., 2017). Heterogeneity in accelerometer measurement and data analysis across existing studies precluded assessment of the influence of data processing decisions and measurement specifications on the final derived profiles. Currently, it is still unclear whether and how physical behavior profiles identified in one sample population could differ from those identified in another population independent of differences attributed to accelerometer data and data processing decisions. It is also unclear how differences in summary metrics and variables derived from accelerometer data, such as MIMS and Euclidean Norm Minus One (ENMO) (Migueles et al., 2019), could have influenced the final profiles identified. The profiling algorithms used for profile analysis (e.g., K-means, latent profile analysis, etc.) may influence the profiles ultimately identified (Brusco et al., 2017). However, it remains unclear to what extent the reported physical behavior profiles depend on the specific algorithm employed. Further research is needed to better understand the advantages and limitations of different algorithms for physical behavior profiling.

## CONCLUSIONS

Data-driven population segmentation analysis is well suited for exploring the complex and potentially unknown interplay among multiple components and aspects of physical behaviors, particularly for investigating how these behaviors could cluster together and synergistically influence health outcomes and indicators. The use of data-driven techniques on wearable accelerometer-measured physical behaviors has generated several hypothesis-generating, data-driven findings that warrant further investigation. Multiple studies have identified data-driven profiles that could be distinguished by the temporal distribution of PA at specific times during a 24-hour period. Joint profiles emerging in data-driven studies indicate that being physically active while simultaneously spending considerable amounts of time in sedentary behaviors may represent a profile present among the adult population. Beyond total sedentary time, patterns of accumulation and interruption of sedentary time may also be linked with health risks and indicators. Although few studies have identified sleep and rest–activity profiles, data-driven findings suggest that bedtime and wake-up patterns and the timing of activity onset may be related to health outcomes.

## Ethics approval and consent to participate

Not applicable.

## Consent for publication

Not applicable.

## Availability of data and materials

No datasets were generated or analysed during the current study.

## Authors’ contributions

VF conceived the idea, conducted the literature search, reviewed the titles and abstracts, and prepared the manuscript draft. EF contributed to assessing the eligibility of articles identified during the title and abstract screening and revised the manuscript.

## Data Availability

All data produced in the present work are contained in the manuscript

## Acknowledgments

None.

