## Supplemental file 1 for "Data-driven profiling of accelerometer-determined physical behaviors and health outcomes in adults: A systematic review"

**Search term**

1. **Pubmed**

("clustering"[Title/Abstract] OR "cluster analysis"[Title/Abstract] OR "profile analysis"[Title/Abstract] OR "data driven"[Title/Abstract] OR "data-driven"[Title/Abstract] OR "person centered"[Title/Abstract] OR "unsupervised"[Title/Abstract])

AND

("sedentary behavior"[Title/Abstract] OR "sedentary behaviour"[Title/Abstract] OR "physical activity"[Title/Abstract] OR "physical activities"[Title/Abstract] OR "sleep"[Title/Abstract] OR "movement behavior"[Title/Abstract] OR "movement behaviour"[Title/Abstract] OR "sleep"[Title/Abstract] OR "activity behavior"[Title/Abstract] OR "activity behaviour"[Title/Abstract] OR "accelerometer"[Title/Abstract] OR "accelerometry"[Title/Abstract] OR "activity monitor"[Title/Abstract])

2. **Web of Science**

("clustering" OR "cluster analysis" OR "profile analysis" OR "data driven" OR "data-driven" OR "person centered" OR "unsupervised")

AND

("sedentary behavior" OR "sedentary behaviour" OR "physical activity" OR "physical activities" OR “sleep” OR "movement behavior" OR "movement behaviour" OR "sleep" OR "activity behavior" OR "activity behaviour" OR "accelerometer" OR “accelerometry” OR "activity monitor")

3. **Scopus**

("clustering" OR "cluster analysis" OR "profile analysis" OR "data driven" OR "data-driven" OR "person centered" OR "unsupervised")

AND

("sedentary behavior" OR "sedentary behaviour" OR "physical activity" OR "physical activities" OR “sleep” OR "movement behavior" OR "movement behaviour" OR "sleep" OR "activity behavior" OR "activity behaviour" OR "accelerometer" OR “accelerometry” OR "activity monitor")

**Table S1:** Device type and details of accelerometer measurement and data processing criteria used in 39 included studies in the review.

| **Study (first author, year)** | **Device** | **Uniaxial or triaxial** | **Unit of measurement** | **Device placement** | **Days monitored** | **Measurement period** | **Number of valid days** | **Valid hours** |
| --- | --- | --- | --- | --- | --- | --- | --- | --- |
| (Yerramalla et al., 2024) | GENEActiv | Triaxial | Raw acceleration | Non-dominant wrist | 9 days | 24-hour | At least 2 weekdays and 2 weekend days | Wear time ≥2/3 of the waking period |
| (Zhang et al., 2024) | ActiGraph GT3X+ | Triaxial | Raw acceleration | Non-dominant wrist | 7 days | 24-hour | 4 days or more | 10 hours or more |
| (Bai, Zhou, et al., 2024) | Axivity AX3 | Triaxial | Raw acceleration | Dominant wrist | 7 days | 24-hour | NR | NR |
| (Fabrizio et al., 2024) | GENEActiv | Triaxial | Raw acceleration | Wrist | 14 days | 24-hour | NR | NR |
| (Chung et al., 2024) | Actiwatch Spectrum | NR | NR | Wrist | 7 days | NR | NR | Valid nights were considered with no more than an hour of “off time” |
| (Shim et al., 2023) | ActiGraph GT3X+ | Triaxial | Raw acceleration | Non-dominant wrist | 7 days | 24-hour | At least 4 days | 16 hour or more |
| (von Rosen et al., 2020) | ActiGraph AM7164 | Uniaxial | CPM | Lower back | 7 days | Awake | At least 1 day | 10 hours or more |
| (Evenson et al., 2017) | ActiGraph AM7164 | Uniaxial | CPM | Hip | 7 days | Awake | 3 days or more | 8 hours or more |
| (Yamamoto et al., 2025) | Lifecorder-  Ex | Uniaxial | NR | NR | 7 days | Awake | 4 days or more | 8 hours or more |
| (Albalak et al., 2023) | Axivity AX3 | Triaxial | Raw acceleration | Dominant wrist | 7 days | 24-hour | Minimum of three days | NR |
| (Bai, Ning, et al., 2024) | Axivity AX3 | Triaxial | Raw acceleration | Dominant wrist | 7 days | 24-hour | NR | NR |
| (Q. Feng et al., 2025) | Axivity AX3 | Triaxial | Raw acceleration | Dominant wrist | 7 days | 24-hour | NR | NR |
| (Bai, Shao, et al., 2024) | Axivity AX3 | Triaxial | Raw acceleration | Dominant wrist | 7 days | 24-hour | NR | NR |
| (Li et al., 2024) | Axivity AX3 | Triaxial | Raw acceleration | Dominant wrist | 7 days | 24-hour | Minimum of 72 hours of data | NR |
| (Reuter et al., 2020) | ActiGraph GT3X+ | Triaxial | Raw acceleration | Hip | 7 days | 24-hour | At least four days | 10 hours or more |
| (Choi et al., 2025) | ActiGraph GT3X+ | Triaxial | Raw acceleration | NR | 7 days | Awake | 3 days or more | 10 hours or more |
| (Gupta et al., 2025) | ActiGraph GT3X+ | Triaxial | Raw acceleration | Right thigh | 4 consecutive days including 2 working days | 24-hour | At least one valid day | At least 4 hours or 75% of the wearer’s average wok and leisure time. |
| (Farrahi et al., 2024) | ActiGraph AM7164 | Uniaxial | CPM | Hip | 7 days | Awake | 4 days or more | 10 hours or more |
| (Leskinen et al., 2023) | activPAL3 | Triaxial | Raw acceleration | Right thigh | 7 days | 24-hour | At least 4 days including one weekend day | Over 10 hours |
| (Ma et al., 2023) | ActiGraph AM7164 | Uniaxial | CPM | Hip | 4 days | Awake | At least 4 days | 10 hours or more |
| (Biswas et al., 2023) | Actical | NR | CPM | Right hip | 7 days | Awake | 4 days or more | 10 hours or more |
| (Yamamoto et al., 2023) | Lifecorder-Ex | Uniaxial | NR | NR | 7 days | Awake | 4 days or more | 8 hours or more |
| (Biswas et al., 2022) | Actical | NR | CPM | Right hip | 7 days | Awake | 4 days or more | 10 hours or more |
| (Farrahi et al., 2022) | Hookie | Triaxial | Raw acceleration | Hip | 14 days | Awake | 7 consecutive days | 10 hours or more |
| (Hibbing et al., 2022) | ActiGraph AM7164 | Uniaxial | CPM | Hip | 7 days | Awake | 4 days or more | More than 10 hours |
| (Aqeel et al., 2021) | ActiGraph AM7164 | Uniaxial | CPM | Hip | 7 days | Awake | One random day from all valid days | 10 hours |
| (Farrahi et al., 2021) | Hookie | Triaxial | Raw acceleration | Hip | 14 days | Awake | 7 consecutive days | 10 hours or more |
| (Matricciani et al., 2021) | GENEActiv | Triaxial | Raw acceleration | Wrist | NR | NR | At least four nights of sleep | Sleep time >200 min and at least one night on Fri-Sat |
| (Gupta et al., 2020) | ActiGraph GT3X+ | Triaxial | Raw acceleration | Right thigh | 4 consecutive days including 2 working days | 24-hour | At least one valid day | At least 4 hours or 75% of the wearer’s average wok and leisure time. |
| (Niemelä et al., 2019) | Polar Active | Uniaxial | MET values | Wrist | 14 days | 24-hour | 7 consecutive days | 10 hours or more |
| (Marschollek, 2016) | ActiGraph AM7164 | Uniaxial | CPM | Hip | 7 days | Awake | NR | NR |
| (P. H. Lee et al., 2013) | ActiGraph AM7164 | Uniaxial | CPM | Hip | 4 days | Awake | 4 days | At least 12 hours |
| (Fukuoka et al., 2018) | Active Style Pro | Triaxial | Raw acceleration | Hip | 7 days | Awake | All 7 days | At least 8 hours |
| (Nawrin et al., 2024) | ActiGraph GT3X+ | Triaxial | Raw acceleration | Wrist | 7 days | 24-hour | 7 days | NR |
| (Smagula et al., 2022) | ActiGraph GT3X+ | Triaxial | Raw acceleration | Non-dominant wrist | 7 days | 24-hour | NR | NR |
| (O’Regan et al., 2021) | ActivPAL 3 | Triaxial | Raw acceleration | Right thigh | 8 days | 24-hour | At least 3 weekdays and 1 weekend | 10 hours or more |
| (Palmberg et al., 2024) | UKK RM42 | Triaxial | Raw acceleration | Dominant thigh | 3-10 days | 24-hour | At least 3 complete days | Complete days with no non-wear |
| (Graves et al., 2021) | ActiGraph GT3X+ | Triaxial | Raw acceleration | Non-dominant wrist | 7 days | 24-hour | Minimum of 3 days | At least 10 hours |
| (Full et al., 2019)* | Actigraph GT3X+ | Triaxial | Raw acceleration | Two devices worn concurrently on hip and wrist | 7 days | Hip (Awake)  Wrist (24-hour) | Hip worn device: 5 days  Wrist worn device: NR | Hip worn device: At least 10 hours  Wrist worn device: NR |
| (Paolillo et al., 2023) | Fitbit Flex 2 | Triaxial | Proprietary | NR | 30 continuous days | 24-hour | NR | NR |

*The hip-worn device was used to measure physical activity during waking hours, while the wrist-worn device was worn continuously for 24 hours and used to obtain sleep parameters. Abbreviations: NR = Not reported; CPM = Counts per minute;

Adapted quality assessment scale used in this study was based on the combination of items from Newcastle-Ottawa Quality Assessment Scale Cohort Studies (Wells et al., 2000) and recommendations by (Montoye et al., 2018) for reporting accelerometer methods.

Four items were selection from Newcastle-Ottawa Quality Assessment Scale for Cohort Studies (Wells et al., 2000) and recommendations by (Montoye et al., 2018) for reporting accelerometer methods. A study can be awarded a maximum of one point for each numbered item within the Selection, Outcome, and Accelerometer information, data processing and interpretation categories. If any of the categories marked with * is selected, the point given to the study for that item is 1.

**Selection**

1) Representativeness of the exposed cohort
a) truly representative *
b) somewhat representative *
c) selected group of users eg nurses, volunteers
d) no description of the derivation of the cohort
2) Ascertainment of exposure
a) secure record (eg surgical records) *
b) structured interview *
c) written self-report
d) no description
3) Demonstration that outcome of interest was not present at start of study (not applicable for cross-sectional studies)
a) yes *
b) no

**Outcome**

4) Assessment of outcome
a) independent blind assessment *
b) record linkage *
c) self report
d) no description

**Accelerometer information, data processing and interpretation**

5) Brand of accelerometer used is specified.
   a. yes *
   b. no
6) Model of accelerometer is specified.
a. yes *
b. no
7) Placement of accelerometer is specified.
a. yes *
b. no
8) Days of protocol is specified.
a. yes *
b. no
9) Non-wear criteria is described.
a. yes *
b. no
10) Hours/minutes needed to be considered a valid day is specified.
a. yes *
b. no
11) Number of valid days needed is specified.
a. yes *
b. no
12) Metrics and meaning of metrics derived from accelerometer signal are clearly specified.
a. yes *
b. no

**Table S2:** Assessment of bias according to items adapted from Newcastle-Ottawa Quality Assessment Scale for Cohort Studies [29] and recommendations by Montoye et al [30].

|  | **Selection** | | | **Outcome** | **Accelerometer information, data processing and interpretation** | | | | | | | |  |
| --- | --- | --- | --- | --- | --- | --- | --- | --- | --- | --- | --- | --- | --- |
| **Study (first author, year)** | **1** | **2** | **3*** | **4** | **5** | **6** | **7** | **8** | **9** | **10** | **11** | **12** | **Score** |
| **Prospective studies (n = 14)** |  |  |  |  |  |  |  |  |  |  |  |  |  |
| (Yerramalla et al., 2024) | 1 | 1 | 1 | 1 | 1 | 1 | 1 | 1 | 1 | 1 | 1 | 1 | 12▲ |
| (Zhang et al., 2024) | 1 | 1 | 1 | 1 | 1 | 1 | 1 | 1 | 1 | 1 | 1 | 1 | 12▲ |
| (Bai, Zhou, et al., 2024) | 1 | 1 | 1 | 1 | 1 | 1 | 1 | 1 | 0 | 0 | 0 | 0 | 8▪ |
| (Fabrizio et al., 2024) | 1 | 1 | 1 | 1 | 1 | 1 | 1 | 1 | 0 | 0 | 0 | 1 | 12▲ |
| (Chung et al., 2024) | 1 | 1 | 1 | 1 | 1 | 1 | 1 | 1 | 0 | 0 | 0 | 1 | 9▪ |
| (Shim et al., 2023) | 1 | 1 | 1 | 1 | 1 | 1 | 1 | 1 | 1 | 1 | 1 | 1 | 12▲ |
| (von Rosen et al., 2020) | 0 | 1 | 1 | 1 | 1 | 1 | 1 | 1 | 1 | 1 | 1 | 1 | 11▲ |
| (Evenson et al., 2017) | 1 | 1 | 1 | 0 | 1 | 1 | 1 | 1 | 1 | 1 | 1 | 1 | 11▲ |
| (Yamamoto et al., 2025) | 0 | 1 | 0 | 1 | 1 | 1 | 0 | 1 | 1 | 1 | 1 | 1 | 9▪ |
| (Albalak et al., 2023) | 1 | 1 | 1 | 1 | 1 | 1 | 1 | 1 | 1 | 0 | 1 | 1 | 11▲ |
| (Bai, Ning, et al., 2024) | 1 | 1 | 1 | 1 | 1 | 1 | 1 | 1 | 1 | 0 | 0 | 1 | 10▲ |
| (Q. Feng et al., 2025) | 1 | 1 | 1 | 1 | 1 | 1 | 1 | 1 | 1 | 0 | 0 | 1 | 10▲ |
| (Bai, Shao, et al., 2024) | 1 | 1 | 1 | 1 | 1 | 1 | 1 | 1 | 1 | 0 | 0 | 1 | 10▲ |
| (Li et al., 2024) | 1 | 1 | 1 | 1 | 1 | 1 | 1 | 1 | 1 | 1 | 1 | 0 | 11▲ |
| (Reuter et al., 2020) | 1 | 1 | 1 | 1 | 1 | 1 | 1 | 1 | 1 | 0 | 1 | 1 | 11▲ |
| **Cross-sectional studies (n = 25)** |  |  |  |  |  |  |  |  |  |  |  |  |  |
| (Choi et al., 2025) | 0 | 1 | - | 1 | 1 | 1 | 0 | 1 | 1 | 1 | 1 | 1 | 9▲ |
| (Gupta et al., 2025) | 0 | 1 | - | 1 | 1 | 1 | 1 | 1 | 1 | 1 | 1 | 1 | 11▲ |
| (Farrahi et al., 2024) | 1 | 1 | - | 1 | 1 | 1 | 1 | 1 | 1 | 1 | 1 | 1 | 11▲ |
| (Leskinen et al., 2023) | 1 | 1 | - | 1 | 1 | 1 | 1 | 1 | 1 | 1 | 1 | 1 | 11▲ |
| (Ma et al., 2023) | 1 | 1 | - | 1 | 1 | 1 | 1 | 1 | 1 | 1 | 1 | 1 | 11▲ |
| (Biswas et al., 2023) | 1 | 1 | - | 1 | 1 | 1 | 1 | 1 | 1 | 1 | 1 | 1 | 11▲ |
| (Yamamoto et al., 2023) | 1 | 1 | - | 1 | 1 | 1 | 0 | 1 | 1 | 1 | 1 | 1 | 10▲ |
| (Biswas et al., 2022) | 1 | 1 | - | 1 | 1 | 1 | 1 | 1 | 1 | 1 | 1 | 1 | 11▲ |
| (Farrahi et al., 2022) | 1 | 1 | - | 1 | 1 | 1 | 1 | 1 | 1 | 1 | 1 | 1 | 11▲ |
| (Hibbing et al., 2022) | 1 | 1 | - | 1 | 1 | 1 | 1 | 1 | 1 | 1 | 1 | 1 | 11▲ |
| (Aqeel et al., 2021) | 1 | 1 | - | 1 | 1 | 1 | 1 | 1 | 1 | 1 | 1 | 1 | 11▲ |
| (Farrahi et al., 2021) | 1 | 1 | - | 1 | 1 | 1 | 1 | 0 | 0 | 1 | 0 | 1 | 9▲ |
| (Matricciani et al., 2021) | 1 | 1 | - | 1 | 1 | 1 | 1 | 1 | 0 | 0 | 0 | 1 | 8▪ |
| (Gupta et al., 2020) | 1 | 1 | - | 1 | 1 | 1 | 1 | 1 | 1 | 1 | 1 | 1 | 11▲ |
| (Niemelä et al., 2019) | 1 | 1 | - | 1 | 1 | 1 | 1 | 1 | 1 | 1 | 1 | 1 | 11▲ |
| (Marschollek, 2016) | 1 | 1 | - | 0 | 1 | 1 | 1 | 1 | 0 | 0 | 0 | 1 | 7▪ |
| (P. H. Lee et al., 2013) | 1 | 1 | - | 0 | 1 | 1 | 1 | 1 | 1 | 1 | 1 | 1 | 10▲ |
| (Fukuoka et al., 2018) | 0 | 1 | - | 1 | 1 | 1 | 1 | 1 | 1 | 1 | 1 | 1 | 10▲ |
| (Nawrin et al., 2024) | 1 | 1 | - | 1 | 1 | 1 | 1 | 1 | 0 | 0 | 1 | 1 | 9▲ |
| (Smagula et al., 2022) | 1 | 1 | - | 1 | 1 | 1 | 1 | 1 | 0 | 0 | 0 | 1 | 8▪ |
| (O’Regan et al., 2021) | 0 | 1 | - | 1 | 1 | 1 | 1 | 1 | 1 | 1 | 1 | 1 | 10▲ |
| (Palmberg et al., 2024) | 0 | 1 | - | 1 | 1 | 1 | 1 | 1 | 1 | 1 | 1 | 1 | 10▲ |
| (Graves et al., 2021) | 0 | 1 | - | 1 | 1 | 1 | 1 | 1 | 1 | 1 | 1 | 1 | 10▲ |
| (Full et al., 2019) | 0 | 1 | - | 1 | 1 | 1 | 1 | 1 | 1 | 0 | 0 | 1 | 8▪ |
| (Paolillo et al., 2023) | 0 | 1 | - | 1 | 1 | 1 | 0 | 1 | 0 | 0 | 0 | 1 | 6▪ |

The 12 criteria used for assessment of bias are shown. The maximum number of points that prospective studies can get is 12. The maximum number of points that prospective studies can get is 11. *Only for prospective studies. ▪, moderate quality; ▲, high quality.
